# DRIPS: Domain Randomisation for Image-based Perivascular space Segmentation

**DOI:** 10.1101/2025.10.22.25337423

**Authors:** Luna Bitar, Mario Díaz, Roberto Duarte Coello, Maria d.C. Valdés-Hernández, Hendrik Mattern, Katja Neumann, Malte Pfister, Coretta Dongmo, Carolin Beck, Trong Huy Mai, Erelle Fuchs, Serena Tang, Duygu Tosun, Bianca Besteher, Tonia Rocktäschel, Philipp A. Reuken, Andreas Stallmach, Nils Opel, Christian Gaser, Martin Walter, Marc Dörner, Philipp Arndt, Daniel Behme, Christiane Piechowiak, Yves Lading, Patrick Müller, Rüdiger Braun-Dullaeus, Sven G. Meuth, Merel M. van der Thiel, Jacobus F. A. Jansen, Eva M. van Heese, Max A. Laansma, the Alzheimer’s Disease Neuroimaging Initiative, Joanna M. Wardlaw, Stefanie Schreiber, Maria Trujillo, Emrah Düzel, Gabriel Ziegler, Jose Bernal

## Abstract

**Background:** MRI-visible perivascular spaces (PVS) are an imaging marker of compromised brain health function. However, robust PVS segmentation remains challenging: conventional methods often require cohort-specific parameter tuning, while supervised deep-learning models depend on extensive manual annotations for training and may fail to generalise to unseen scanners, protocols, and clinical populations.

**Methods:** We introduce DRIPS (Domain Randomisation for Image-based PVS Segmentation), a fully automated deep-learning framework trained exclusively on synthetic data. By learning PVS representations from a plethora of simulated cases, DRIPS transfers to real-world MRI without using manually labelled PVS data from real participants. We evaluated DRIPS in 185 fully manually annotated MRI scans from eight independent cohorts encompassing diverse clinical populations and acquisition settings, including 3T and 7T imaging, T1-weighted and T2-weighted sequences, and isotropic and anisotropic resolutions. DRIPS was compared with five previously published methods.

**Results:** Across cohorts, DRIPS significantly outperformed the second-best method for both area under the precision-recall curve (AUPRC) and centreline Dice similarity coefficient (clDice), with mean gains of at least 0.1: 0.146 for basal ganglia (BG) AUPRC (P = 0.008), 0.125 for centrum semiovale (CSO) AUPRC (P = 0.008), 0.112 for BG clDice (P = 0.016), and 0.186 for CSO clDice (P = 0.008). Importantly, its performance was not associated with white matter hyperintensity burden.

**Conclusions:** DRIPS enables accurate, fully automated PVS segmentation across heterogeneous imaging settings without the need for real PVS annotations for training.

## 1 Introduction

Perivascular spaces (PVS) are a brain-wide network of conduits around arterioles, capillaries, and venules in the brain (Gouveia-Freitas and Bastos-Leite, 2021; Wardlaw et al., 2020) that facilitate cerebrospinal fluid (CSF) circulation (Babin et al., 2025; Cengija et al., 2026; Hirschler et al., 2025; Wardlaw et al., 2020, 2009; Yamamoto et al., 2024)—a function that underlies metabolic and neurotoxic waste product clearance (Braun and Iliff, 2020; Hablitz and Nedergaard, 2021; Iliff et al., 2014, 2012; Mestre et al., 2018; Rasmussen et al., 2018; Wåhlin et al., 2026; Wardlaw et al., 2020). These spaces are dynamic, with the capacity to shrink and enlarge, at times reaching a calibre that renders them visible and invisible *in vivo* on magnetic resonance imaging (MRI) (Einspänner et al., 2026; Kern et al., 2023; Kim et al., 2023; Libecap et al., 2023; Lynch et al., 2023; Menze et al., 2024; Morozova et al., 2024; Schipper et al., 2025; Sommer et al., 2025; Vikner et al., 2022; Wuerfel et al., 2008). Although PVS enlargement is inherently a multifactorial process, its chronicity is widely recognised as pathological (Bown et al., 2022; Francis et al., 2019; Okar et al., 2023; Wardlaw et al., 2020; Waymont et al., 2024) and is increasingly discussed in the context of ageing, brain atrophy, cerebrovascular dysfunction, proteinopathies, impaired brain waste clearance, and systemic and neuro-inflammation (Arndt et al., 2026; Babin et al., 2025; Francis et al., 2019; Ineichen et al., 2022; Okar et al., 2023; Schreiber et al., 2023; Wardlaw et al., 2020; Waymont et al., 2024). Importantly, it has been hypothesised—and in multiple instances shown—that these structural alterations are associated with downstream cognitive impairment (Barisano et al., 2025; Chappell et al., 2025; Francis et al., 2019; Javierre-Petit et al., 2020; Kohno et al., 2026; Paradise et al., 2021; Passiak et al., 2019; Sepehrband et al., 2021; Sinclair et al., 2026; Tang et al., 2026a; Wang et al., 2022).

The growing recognition of PVS as a non-invasive imaging marker of compromised brain health function has prompted the development and large-scale deployment of computational methods for their quantification and monitoring (Smith et al., 2019; Waymont et al., 2024). The literature broadly describes two strategies for MRI-based PVS segmentation: non-machine learning and machine learning-based methods (Waymont et al., 2024). Non-machine learning-based methods generally comprise semi-automated techniques, such as intensity thresholding methods, Hessian-based filters, and path-operator-based morphological filters (Ballerini et al., 2020, 2018; Barisano et al., 2025; Barnes et al., 2022; Bernal et al., 2021b, 2020; Boespflug et al., 2018; Duarte Coello et al., 2024; Gibson et al., 2026; Menze et al., 2024; Schwartz et al., 2019; Valdés Hernández et al., 2024). Intensity thresholding segments PVS by exploiting their characteristic CSF-like signal intensity, classifying voxels whose intensity falls within manually defined ranges. Hessian-based filters detect tubular structures by performing eigenvalue decomposition of the Hessian matrix at each voxel, allowing the characterisation of local curvature and the identification of elongated, tubular structures. Path-operator-based morphological filters detect tubular structures by probing the image along multiple orientations and quantifying morphological path-opening responses, as tubular structures exhibit strong responses along their principal orientation but weaker responses in other directions. These methods often leverage multimodal information to distinguish PVS from other concomitant lesions, such as white matter hyperintensities (WMH). They also offer high sensitivity (Bernal et al., 2022)—a double-edged sword that often requires careful parameter tuning and post-processing to minimise false positives (Ballerini et al., 2018; Bernal et al., 2022, 2020; Gibson et al., 2026; Valdés Hernández et al., 2024).

Machine learning methods, on the other hand, leverage supervised learning (Boutinaud et al., 2021; Cai et al., 2024; Chai et al., 2025; Dubost et al., 2019a, 2019b; González-Castro et al., 2016; Hayashi et al., 2026; Hou et al., 2017; Lian et al., 2018; Park et al., 2016; Pham et al., 2026; Rashid et al., 2023; Sinclair et al., 2026; Tang et al., 2026a; Zhang et al., 2017). Within this category, deep learning has emerged as the most widely adopted method (Waymont et al., 2024). Its main advantage is its potential to overcome the limitations of non-machine-learning approaches, provided it is trained on sufficiently large, diverse, and well-annotated datasets. In practice, however, such datasets are rarely available (Sudre et al., 2024) and are likely to remain so since their creation requires labour-intensive, resource-intensive expert annotation. In such annotation-scarce settings, models tend to exploit dataset-specific shortcut features that fail to transfer to unseen data. This problem was evident in the MICCAI 2024 PVS Segmentation Challenge (Wu et al., 2026) and the DoRa-PVS Segmentation Challenge 2026 (Dash et al., 2027), where models experienced severe performance degradation on data acquired with unseen protocols and scanners (see Fig. 7 in (Wu et al., 2026) and Fig. 1 in (Dash et al., 2027)). The central concern is that, if insufficiently characterised, these quantification errors may be large enough to obscure expected biological associations of PVS (Tang et al., 2026b).

**Figure 1.**
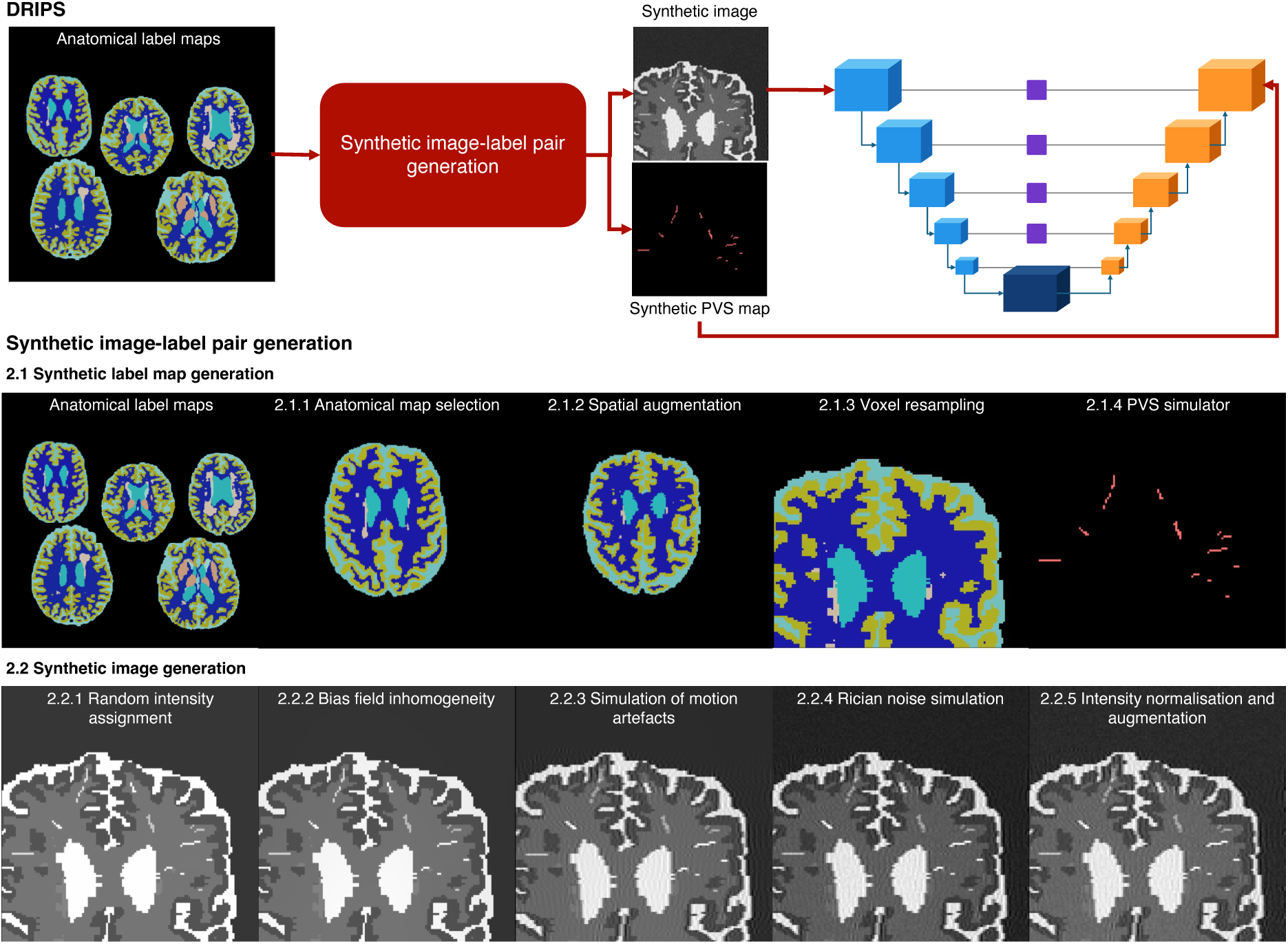
DRIPS is a domain randomisation framework for accurate, fully automated PVS segmentation across heterogeneous imaging settings and clinical populations. DRIPS generates PVS masks from location, shape, and orientation priors inspired by radiological standards, and combines them with anatomical label maps to create synthetic images with variable contrast, resolution, artefacts, and co-occurring pathology. These synthetic image-label pairs are used to train segmentation networks. At inference, DRIPS automatically produces native-space voxel-wise PVS probability maps and segmentations.

Given that large, heterogeneous annotated datasets are likely to remain scarce, alternative strategies are needed to enable deep learning models to generalise beyond their training data in the absence of real annotated data. A promising approach is synthetic training, particularly domain randomisation, in which models are trained on synthetic image-label pairs generated with randomised nuisance factors—e.g., intensity distributions, contrast, noise, resolution, and artefacts. By deliberately introducing variability during case generation, domain randomisation aims to train models that are robust to differences in scanners, acquisition protocols, image quality, and study populations. SynthSeg successfully illustrates the potential of domain randomisation: models trained exclusively on its generated cases can segment brain structures in real MRI across diverse sequences and modalities without retraining (Billot et al., 2023a). Since its introduction in the early 2020s, domain randomisation has been fruitfully applied to a variety of tasks, including skull stripping (Hoopes et al., 2022); segmentation of brain structures (Billot et al., 2023a, 2023b; Valabregue et al., 2026), WMH (Laso et al., 2024), and stroke lesions (Chalcroft et al., 2025); as well as super-resolution (Iglesias et al., 2023) and image registration (Hoffmann et al., 2024).

In this paper, we introduce DRIPS (Domain Randomisation for Image-based PVS Segmentation), the first domain randomisation framework for fully automated PVS segmentation across heterogeneous imaging settings and clinical populations. In contrast to domain randomisation approaches that rely on real target-structure segmentations to generate training cases, DRIPS procedurally generates PVS masks from location, shape, and orientation priors inspired by radiological standards. We evaluated DRIPS in a challenging, heterogeneous setting comprising 185 manually annotated cases with isotropic and anisotropic T1- and T2-weighted MRI acquired at 3 T and 7 T. Cases included cognitively normal individuals and participants with post-COVID syndrome, hypertensive arteriopathy, cerebral amyloid angiopathy, heart failure, mild cognitive impairment, Alzheimer’s disease, Parkinson’s disease, and Lewy body dementia. DRIPS consistently generally outperformed both non-machine-learning and deep-learning comparators and its performance was stable across WMH-burden levels. Together, these findings indicate that DRIPS can learn domain-transferable PVS representations despite training without real images or real manually annotated PVS data.

## 2 DRIPS

DRIPS’s pipeline consists of the following stages (Figure 1): generation of anatomical label maps containing simulated PVS; synthesis of images conditioned on these label maps; training of a deep learning model for binary PVS segmentation; and the inference procedure. Each stage is described in detail in the following sections. *The final parameter values are reported in the Supplementary Material*.

### 2.1 Synthetic label map generation

The first stage generated label maps containing simulated PVS.

#### 2.1.1 Anatomical map selection

The first stage begins by randomly selecting an anatomical label map

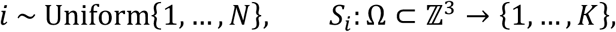

where {*S_n_*}*^N^n*=1 is the finite set of source anatomical label maps. Here, Ω denotes the discrete voxel lattice and *K* the number of anatomical classes before adding the dedicated PVS label. Each map *S̃_i_* assigns a label from {1,…,*K*} to every voxel location **x** = (*x*_1_, *x*_2_, *x*_3_) ∈ Ω. The maps are defined on an isotropic grid with voxel-size vector

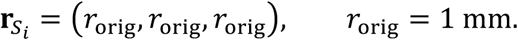

The initial generation label set comprised background, third and fourth ventricles, brainstem, cerebral white matter, cerebral cortical grey matter including hippocampus and amygdala, lateral ventricles, inferior lateral ventricles, cerebellar grey matter, cerebellar white matter, deep grey matter structures, WMH, ventral diencephalon, and extraventricular CSF (eCSF).

The WMH label is important because WMH are a well-documented source of errors in PVS segmentation (Bernal et al., 2022; Waymont et al., 2024). By explicitly modelling WMH during synthetic data generation, we exposed the segmentation model to this key confounder during training and encouraged it to distinguish PVS from WMH.

#### 2.1.2 Spatial augmentation

To increase anatomical variability, each label map *S̃_i_* was spatially augmented using a randomly sampled rigid transformation, *φ*_rigid_. The transformation was applied with nearest-neighbour interpolation to preserve the discreteness of anatomical labels, yielding

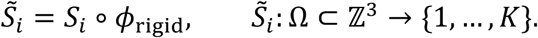

The rigid transformation was parameterised by rotations about and translations along the three spatial axes:

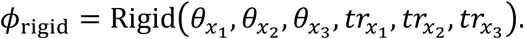

For each spatial dimension *x_j_*, rotation angles and translations were sampled independently from uniform distributions:

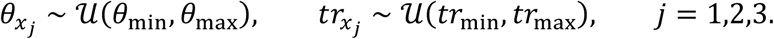

#### 2.1.3 Voxel resampling

Although not inherently a requirement, deep learning-based segmentation frameworks often resample input volumes to an isotropic resolution before inference and subsequently warp the predicted segmentations back to native space (Boutinaud et al., 2021; Pham et al., 2026). This approach is often acceptable for large anatomical structures, whose appearance and boundaries remain relatively stable after interpolation. However, it is poorly suited to PVS, which are small, thin, and often only a few voxels wide. Up- or down-sampling can blur, fragment, merge, or eliminate visible PVS, altering the very morphological and intensity cues required for their detection. Resampling predictions back to native space introduces a further interpolation step that can reduce voxel-wise correspondence and spatial fidelity.

In DRIPS, the segmentation model operates directly in native space. During synthetic-image generation, the pipeline stochastically perturbs the voxel size of anatomical label maps, producing training images that can span both isotropic and anisotropic acquisitions. This voxel-resolution augmentation seeks to expose the model to the resolution-dependent changes in PVS appearance, visibility, and morphology that occur across MRI protocols. The intention is that the model learns to accommodate heterogeneous native resolutions without resampling real images or predictions at inference, preserving the native geometry and voxel-wise localisation of PVS.

Formally, a target voxel-size vector **r**_res_ was stochastically sampled for each transformed label map *S̃_i_*. First, it was determined whether the target resolution was isotropic or anisotropic:

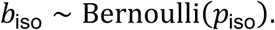

If *b*_iso_ = 1, an isotropic voxel size was sampled

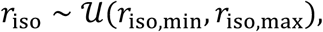

and the target voxel-size vector was defined as

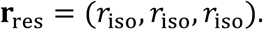

Otherwise, if *b*_iso_ = 0, through-plane and in-plane voxel sizes were independently sampled:

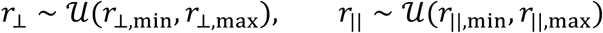

A through-plane axis, *a*∼Uniform{*x*_1_, *x*_2_, *x*_3_}, was then selected uniformly at random, and the target voxel-size vector was defined as

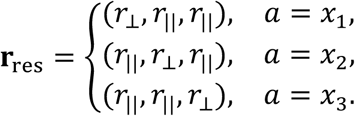

### Resampling and lattice consistency

The resampling operator ℛ**_r_**_res_was applied to *S̃_i_*, yielding

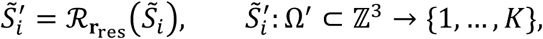

defined on an updated voxel lattice Ω^′^ with voxel size **r**_res_.

Since Ω^′^ may differ from the original lattice Ω in cardinality, a cropping or zero-padding operator ℳ_Ω_ is applied to enforce consistent dimensions. The overall operation can therefore be written compactly as

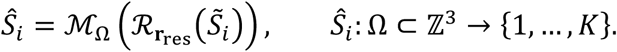

#### 2.1.4 PVS simulator

The stochastic PVS simulator *G*_PVS_ takes an anatomical label map *S̃_i_* and its voxel spacing **r***S̃_i_* =(*r_x_*_1_,*r_x_*_2_,*r_x_*_3_) as input and produces an augmented anatomical label map

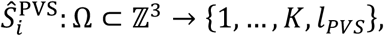

where *l*_PVS_ denotes the dedicated label assigned to simulated PVS.

### Identification of reference regions

Let ℓ_vent_, ℓ_eCSF_, ℓ_brain_, and ℓ_domain_ ⊂ {1,…,*K*} denote the label sets corresponding to ventricular CSF, eCSF, brain tissue, and the admissible PVS domain, respectively. For any label set *l* ⊂ {1,…,*K*}, the corresponding binary mask was defined as

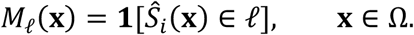

The ventricular, eCSF, brain, and PVS domain masks were therefore given by

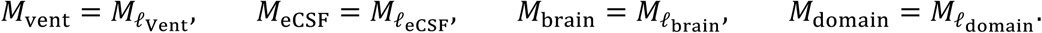

The ventricular and eCSF masks serve as anatomical reference for defining simulated PVS trajectories, while *M*_domain_ restricts PVS generation to plausible brain regions.

### Sampling valid starting locations

Let

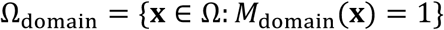

denote the set of valid seed locations. The maximum number of simulated PVS is sampled as

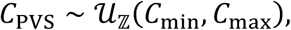

and seed points are sampled uniformly from Ω_domain_,

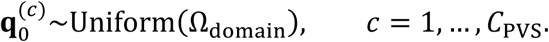

### Vector fields defining trajectories and orientations

PVS exhibit a preferential orientation towards the lateral ventricles: those around lenticulostriate arteries follow their superomedial course from the anterior perforated substance into the dorsal basal ganglia towards the lateral ventricular wall, whereas PVS surrounding medullary arteries course centripetally through the hemispheric white matter in the direction of the lateral ventricles (Duering et al., 2023; Marinković et al., 2001; Wardlaw et al., 2020) (see Figure 2).

**Figure 2.**
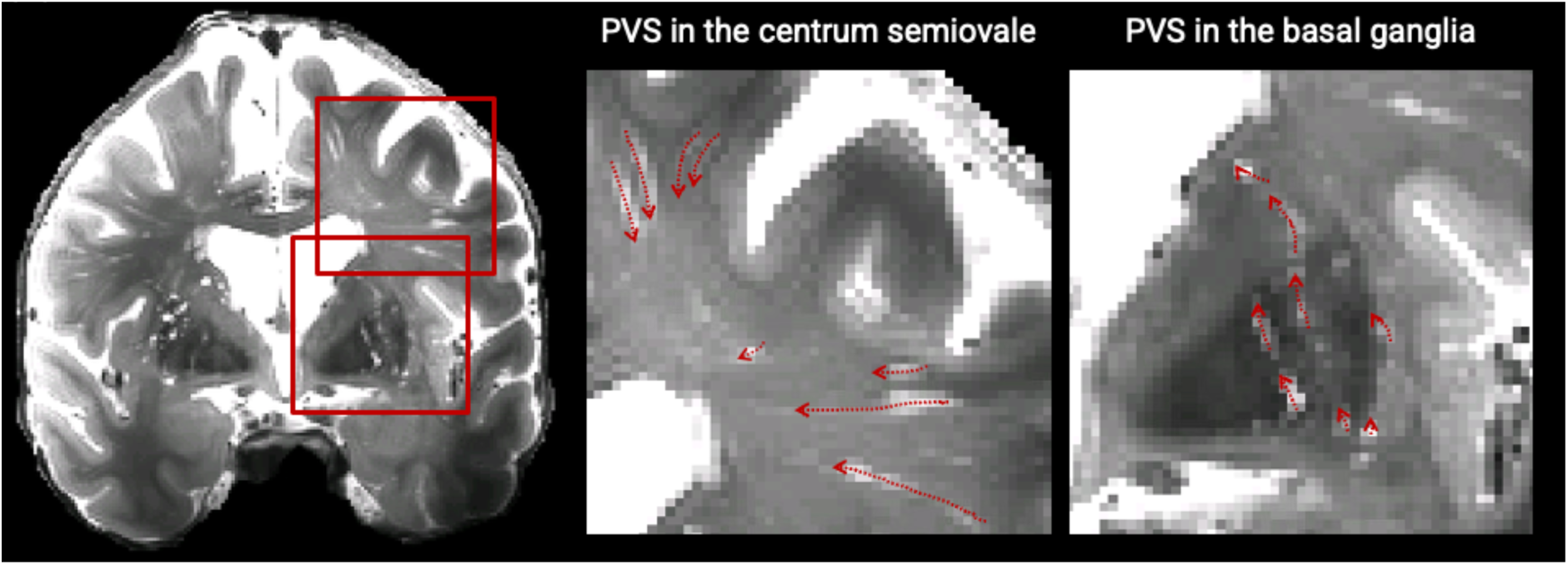
The characteristic orientation of PVS along the lenticulostriate and medullary arteries, often extending towards the lateral ventricles. Note that this description is an over-simplification of the complex three-dimensional orientations of real PVS.

To encode this radiological prior, the simulator uses the ventricles and eCSF as anatomical reference compartments to define simulated PVS trajectories. First, anisotropic Euclidean distance transforms to the ventricular and eCSF compartments were computed:

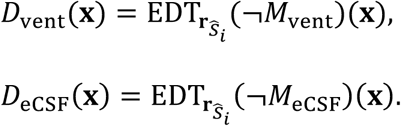

The use of anisotropic distance transforms ensures that distances were measured in physical space, accounting for the voxel spacing of the current synthetic image. The ventricular distance field defines an attraction field towards the ventricles

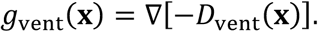

whereas the eCSF distance field defines a repulsion field away from eCSF spaces,

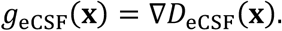

The two components were then combined to obtain a composite guidance field:

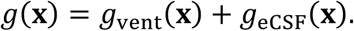

This field encourages simulated PVS to follow trajectories directed towards the ventricles while remaining away from eCSF. To restrict propagation to brain tissue and encode direction rather than speed, the field is masked by the brain mask and normalised to unit magnitude:

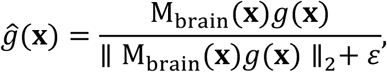

where *ε* > 0 ensures numerical stability. Normalisation ensures that the field encodes directional information rather than magnitude, such that the propagation step size is governed explicitly during path generation rather than by local variations in the distance field gradient.

### Path generation

For each seed point **q**^(*c*)^_0_, a number of propagation steps was sampled from an integer range

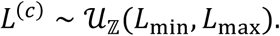

Starting from **q**^(*c*)^_0_, a trajectory was generated by forward Euler integration along the local unit guidance field. At iteration *t*, the guidance direction was perturbed with isotropic Gaussian noise *ĝ_t_*. The updated position was then calculated as

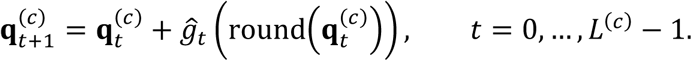

Propagation was terminated if the updated position left the image domain, fell outside the PVS domain, or reached the lateral ventricles.

### Diameter and rasterisation

Consecutive trajectory positions were connected to form a continuous centreline, which was rasterised into a connected voxel chain,

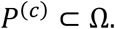

Rather than applying a discrete voxel-width dilation, each centreline was thickened in physical space using a spherical radius *ρ*^(*c*)^ ∼ U_ℤ_(*ρ*_min_, *ρ*_max_). The resulting PVS tube was defined as

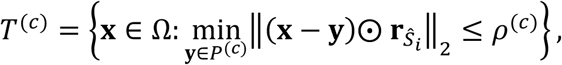

where ⨀ denotes element-wise multiplication. This operation produced a union of capsule-shaped segments with rounded endpoints. Consequently, PVS cross-sections were circular in physical space and were represented appropriately in voxel space for both isotropic and anisotropic images. A candidate tube was added to the PVS mask only if it did not overlap a previously accepted tube:

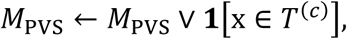

provided that

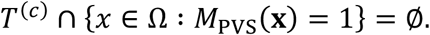

Otherwise, the candidate PVS was discarded to prevent clustering.

### Anatomical constraints and label map update

The final PVS mask is constrained to the domain:

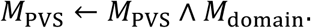

The augmented anatomical label map is then defined as

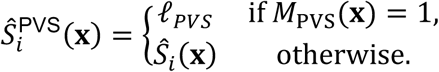

### 2.2 Synthetic image generation

The second stage generated a synthetic image

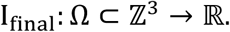

from the anatomically augmented label map containing simulated PVS, *S̃_i_*^PVS^.

#### 2.2.1 Random intensity assignment and partial volume effects

The image synthesis stage began by assigning random intensity values to each anatomical label. Random intensity assignment was therefore constrained to preserve the expected relative PVS intensity while retaining variability in the contrast of other tissues. Specifically, *K* + 1 intensity values were independently sampled and sorted:

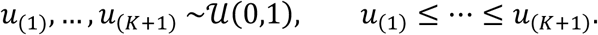

The highest sampled intensity was reserved for CSF and PVS,

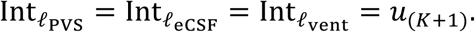

The remaining values were randomly assigned to the non-CSF anatomical labels, subject to the predefined rank constraints for high-intensity tissue classes. The initial synthetic image was constructed voxel-wise as

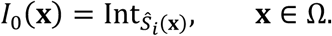

To simulate partial-volume effects, the binary PVS mask was separated into *C*_PVS_ connected components. Each component M^(*c*)^_PVS_ was smoothed with a Gaussian kernel, yielding a soft mask *M̃^(c)^*_PVS_. The kernel width was set to zero along the thick-slice direction for strongly anisotropic images. A component-specific contrast coefficient was sampled as

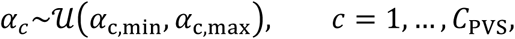

The image was then updated as

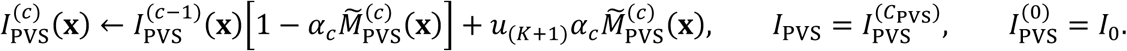

#### 2.2.2 Bias field inhomogeneity

To simulate smooth multiplicative bias-field inhomogeneity, a low-resolution Gaussian random field was sampled on a 4 × 4 × 4 grid (Billot et al., 2023a):

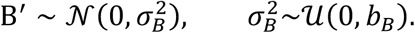

The field was then upsampled to the full image lattice Ω using trilinear interpolation, yielding a smooth bias field *B*: Ω → ℝ. The synthetic image was modulated multiplicatively as

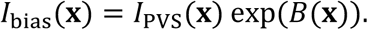

#### 2.2.3 Simulation of motion artefacts

Motion artefacts represent a frequent source of MRI image degradation and can substantially impair the visualisation and quantification of fine anatomical structures such as PVS (Bernal et al., 2022). PVS are particularly vulnerable to confusion with motion-induced streaking artefacts, emphasising the importance of explicitly accounting for them.

Motion artefacts were simulated in *k*-space to model rotational head motion occurring during image acquisition (Bernal et al., 2022, 2021a; Shaw et al., 2020). Two motion-corrupted intermediate volumes were generated as

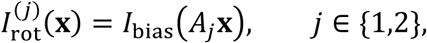

where each *A_j_* ∈ ℝ^3×3^ is a 3D rotation matrix with rotation angles sampled uniformly from *U*(*R_θ_*_min_, *R_θ_*_max_) about randomly selected spatial axes.

Let ℱ and ℱ^-1^ denote the centred forward and inverse 3D Fourier transforms, respectively. The corresponding k-space representations are

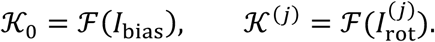

A composite k-space *K̃* was then constructed by retaining a random proportion *pr* ∼ *U*(*pr*_min_, *pr*_max_) of frequency lines from K_0_, while replacing the remaining lines with lines from K^(1)^ and K^(2)^along a randomly selected acquisition axis. This strategy mimics rotational motion occurring intermittently during *k*-space acquisition.

The amplitude and relative timing of the simulated rotations determine the severity and visual appearance of the resulting motion artefacts.

#### 2.2.4 Rician noise simulation

Rician noise in MRI originates from additive white Gaussian noise in *k*-space affecting the real and imaginary signal components, and emerges after inverse Fourier transformation and magnitude reconstruction (Gudbjartsson and Patz, 1995).

Complex white Gaussian noise was added to *K̃* as

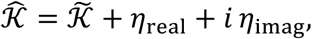

where *η*_real_, *η*_imag_ ∼ N(0, σ^2^_noise_) are independent and identically distributed. The noise level was defined as

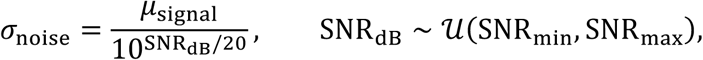

where *μ*_signal_ denotes the mean magnitude of the signal.

The final image with motion artefacts and Rician noise was obtained via inverse Fourier transform and magnitude reconstruction

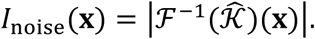

#### 2.2.5 Intensity normalisation and augmentation

A random gamma-intensity transformation was applied to each synthetic image to increase or decrease variation in global image contrast (Billot et al., 2023a). The image was first min-max normalised to [0, 1], after which a gamma value was sampled uniformly from

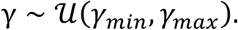

The transformed image was defined as

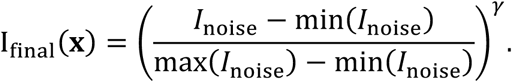

Gamma values below one increased the relative brightness of low-intensity voxels, whereas values above one darkened them.

#### 2.2.6 Pair of synthetic image and label map

Figure 3 shows two examples of synthetic images *I*_final_ with their corresponding binary PVS masks *M*_PVS_, highlighting the variability in anatomy and intensity induced by DRIPS. These types of image-mask pairs were used as input-output data for training segmentation models.

**Figure 3.**
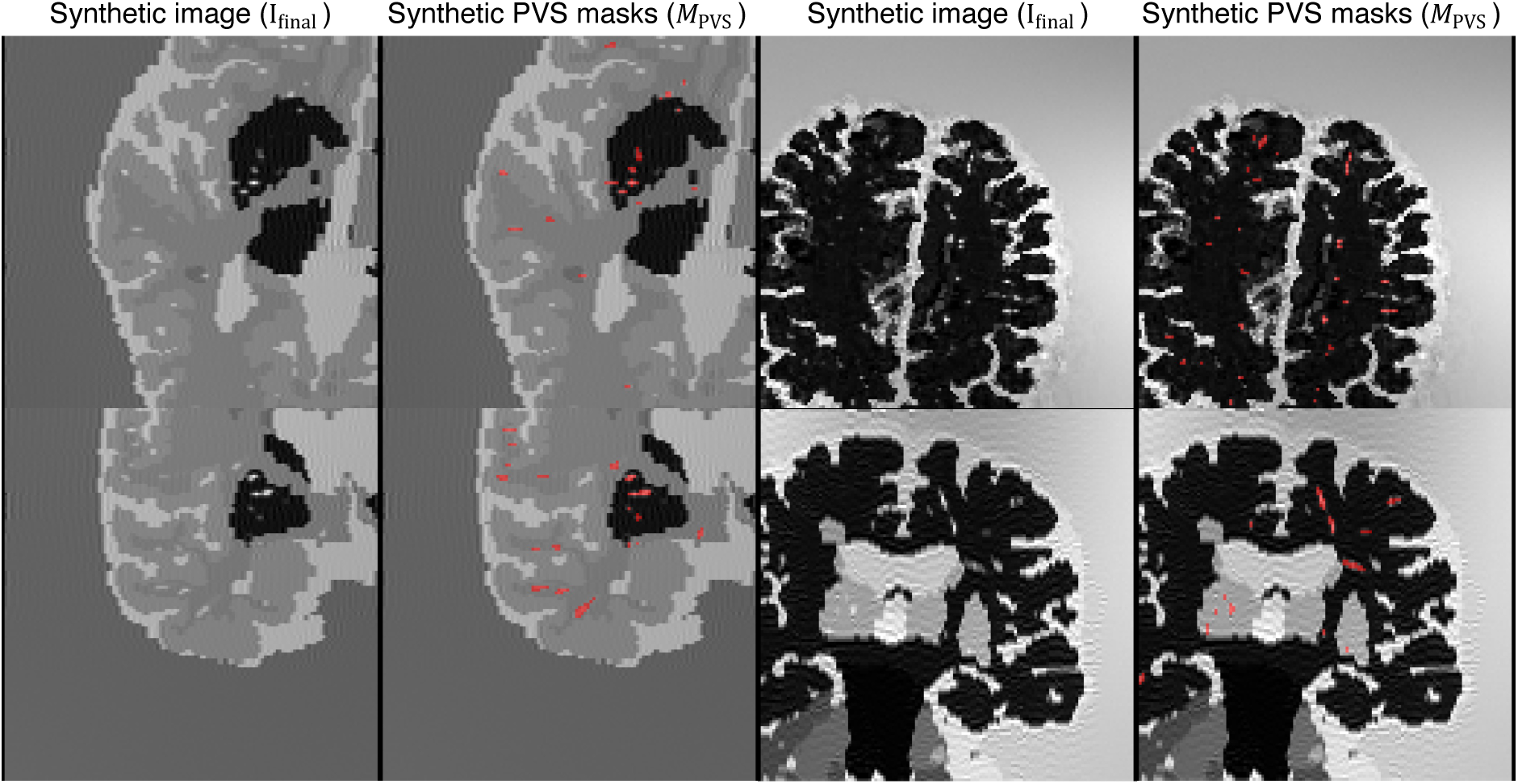
Examples of synthetic images and their corresponding PVS maps. The top row shows axial views, and the bottom row shows coronal views. PVS appear in red in the synthetic PVS masks.

### 2.3 Segmentation model training

#### 2.3.1 Anatomical label maps

The anatomical priors used in the procedural image generation pipeline were derived from T1-weighted and FLAIR MRI scans of 839 individuals from ADNI and the CSVD Magdeburg cohort (Arndt et al., 2025).

Their construction involved three steps. First, we segmented brain structures from T1-weighted images using SynthSeg (Billot et al., 2023a) and WMH from FLAIR images using LST-AI (Wiltgen et al., 2024). Second, we explicitly integrated WMH voxels identified by LST-AI into the corresponding structural compartments of the SynthSeg-derived label maps. Third, if needed, we resampled all anatomical label maps to an isotropic resolution of 1 mm³ using a nearest neighbour interpolator to standardise voxel geometry across subjects.

Data used in the preparation of this article were obtained from the Alzheimer’s Disease Neuroimaging Initiative (ADNI) database (adni.loni.usc.edu). The ADNI was launched in 2003 as a public-private partnership, led by Principal Investigator Michael W. Weiner, MD. The primary goal of ADNI has been to test whether serial magnetic resonance imaging (MRI), positron emission tomography (PET), other biological markers, and clinical and neuropsychological assessment can be combined to measure the progression of mild cognitive impairment (MCI) and early Alzheimer’s disease (AD). Each participating site obtained institutional review board (IRB) approval, and participants provided written informed consent. Detailed inclusion, exclusion, and diagnostic criteria are published elsewhere (Jack et al., 2024).

#### 2.3.2 Segmentation model

The segmentation model takes an image *I*: Ω ⊂ ℤ^3^ → ℝ and its voxel size vector **r**_I_ = (*r_x_*_1_, *r_x_*_2_, *r_x_*_3_) as input and produces a PVS probability map *p*: Ω → [0,1]. During training, *I* corresponds to a synthetic image *I*_final_ generated as described above. At inference, it corresponds to a real image, preprocessed such that PVS are hyperintense.

The model combines a fixed Laplacian front-end, a three-dimensional Attention U-Net, and a convolutional refinement module. First, to highlight structures compatible with the expected intensity profile of hyperintense PVS, we computed a fixed three-dimensional Laplacian representation, Δ**_r_***_I_I*, using the standard second-order finite-difference approximation adjusted for the voxel size of the input image **r**_I_. Because PVS were modelled as locally hyperintense structures, we inverted, normalised, and rectified the Laplacian response:

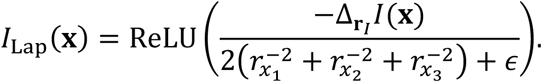

The original image and the Laplacian representation were concatenated channel-wise, *I*_cat_ = [*I*, *I*_Lap_], and provided as input to an Attention U-Net (Oktay et al., 2018), implemented using MONAI. The network comprised five resolution levels with 32, 64, 128, 256, and 512 channels, respectively, and four successive 2 × 2 × 2 downsampling operations. All convolutional and up-convolutional kernels had size 3 × 3 × 3. Attention gates in the decoder selected encoder features relevant to PVS segmentation before their fusion through skip connections. The Attention U-Net produced an initial voxel-wise logit map

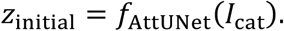

To refine this initial prediction, the original image, Laplacian map, and initial response map were concatenated and processed by two 3 × 3 × 3 convolutional layers. The first layer produced 16 feature maps and was followed by a LeakyReLU activation with negative slope 0.1. The second layer produced the final voxel-wise response. The refinement module yielded a learned voxel-wise probability map

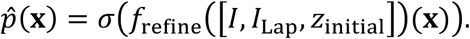

The final prediction combined this learned response with the Laplacian-derived local image prior,

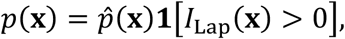

retaining positive responses only at locations with the expected intensity polarity.

#### 2.3.3 Training setup

We trained the segmentation model for 100 epochs using 1000 image-mask pairs (*I*_final_, *M*_PVS_) generated on the fly in each epoch, as described in Sections 2.1 and 2.2. We used an equally weighted composite loss comprising focal, Tversky, and Skeleton Recall Loss terms (Kirchhoff et al., 2024; Lin et al., 2017; Sadegh et al., 2017):

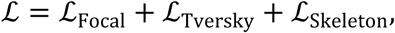

to handle complementary failure modes: focal loss reduces the dominance of easily classified background voxels, Tversky loss prioritises recovery of subtle PVS, and Skeleton Recall Loss encourages detection of their centreline regions.

Let *p*(**x**) ∈ [0, 1] denote the predicted PVS probability and *M*_PVS_(*x*) ∈ {0, 1} the reference label at voxel *x*. The focal loss was defined as

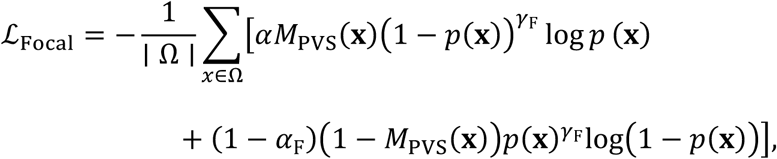

where *α*_F_ up-weights PVS voxels relative to the substantially more prevalent background voxels. The focusing parameter, *γ*_F_, down-weights well-classified voxels, concentrating learning on difficult PVS and background examples.

The Tversky loss was defined as

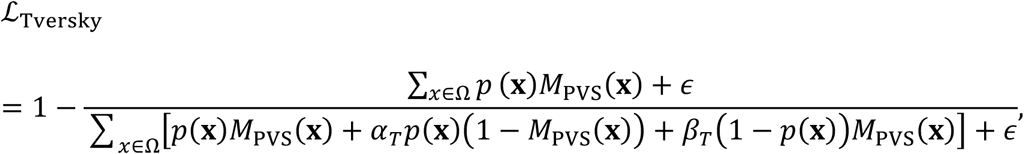

where *α_T_* weights false-positive predictions and *β_T_* weights false-negative predictions.

Finally, we computed the skeleton recall loss using the skeleton of the reference PVS mask, *S*_PVS_,

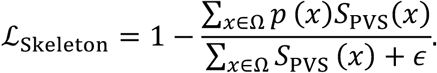

The aim of introducing this term was to encourage accurate segmentation of PVS centrelines and favour continuous rather than fragmented PVS delineations.

We optimised the model using AdamW (Loshchilov and Hutter, 2019), with an initial learning rate of 1×10^-4^ and weight decay of 1×10^−5^. The learning rate followed a cosine-annealing schedule over 100 epochs, decreasing to a minimum value of 1×10^−6^. Training required approximately 30 minutes per epoch and was completed in around 50 hours on an NVIDIA A100 Tensor Core GPU.

### 2.4 Inference

During inference, we used SynthSeg to obtain white matter and CSF masks from each input image. We then compared the intensities within these tissues to infer the CSF-to-white-matter contrast polarity, which determines the expected PVS polarity because PVS are filled with CSF. If white matter was brighter than CSF, PVS were expected to be hypointense relative to white matter. We therefore inverted the image intensities before normalisation. Otherwise, the image was left unchanged. The resulting image was min-max normalised. We extracted its native voxel-spacing vector from the image header and supplied it, together with the normalised image, to the segmentation model. No spatial resampling was performed.

## 3 Evaluation

We evaluated DRIPS in the DoRA-PVS Challenge 2026, a standardised multicohort benchmark designed to assess the generalisability of automated PVS segmentation across heterogeneous MRI datasets (Dash et al., 2027).

### 3.1 Test cohorts and ground truth

The DoRA-PVS Challenge 2026 has eight cohorts with fully manual PVS delineations: REMIND, PCB (Besteher et al., 2022), ADNI-3 (Tang et al., 2026b), GlyM (van der Thiel et al., 2024), EBBIVD, HIM-Study (DRKS00031583) (Müller et al., 2024), MD-DARS (Neumann et al., 2022), and BB-DARS. Their characteristics are summarised in Table 1.

**Table 1.** Imaging protocols and PVS and WMH burden across the DoRA-PVS Challenge 2026 cohorts with fully manual PVS annotations. This table summarises the imaging sequences, acquisition parameters, and scanner characteristics used for manual PVS segmentation in REMIND, PCB, ADNI-3, GlyM, EBBIVD, the HIM Study, MD-DARS, and BB-DARS. PVS and WMH burden are reported separately for the basal ganglia and centrum semiovale regions of interest (BG ROI and CSO ROI) as median absolute and fractional volumes, with interquartile ranges in brackets. Fractional volume denotes the proportion of the regional volume occupied by PVS or WMH.

|  | REMIND | PCB | ADNI | GlyM | EBBIVD | HIM-Study | MD-DARS | BB-DARS |
| --- | --- | --- | --- | --- | --- | --- | --- | --- |
| N | 5 | 42 | 60 | 5 | 18 | 39 | 6 | 10 |
| Clinical groups | Parkinson's disease<br>Dementia with Lewy<br>Bodies | Normal cognition<br>post-COVID<br>syndrome | Normal cognition<br>Mild cognitive<br>impairment<br>Alzheimer's disease | Individuals spanning<br>the Alzheimer's<br>disease continuum | Hypertensive<br>arteriopathy<br>Cerebral amyloid<br>angiopathy | Individuals with heart<br>failure with preserved<br>ejection fraction | Individuals spanning<br>the Alzheimer's<br>disease continuum | Healthy controls<br>Alzheimer's disease<br>Cerebral amyloid<br>angiopathy |
| Imaging setup |  |  |  |  |  |  |  |  |
| Sequence for PVS<br>segmentation | 3D MP2RAGEME<br>3D MP2RAGE | 3D T1w MPRAGE | 3D T1w MPRAGE | 3D T2w TSE | 2D T2w TSE | 2D T2w TSE | 2D T2w TSE | 2D T2w TSE |
| Key parameters | MP2RAGEME:<br>TR: 6.2/31 ms<br>TE: 3.0 ms<br>MP2RAGE:<br>TR: 6.2 ms<br>TE: 2.5 ms | TE = 2.22 ms<br>TR = 2400 ms<br>FA = 9° | TE = min full<br>TR = 2300 ms<br>TI = 900 ms<br>FA = 9° | Before scanner<br>upgrade:<br>TE1 = 204 ms, TE2 =<br>561 ms, TR: 5730 ms<br>After scanner<br>upgrade:<br>TE1 = 204 ms, TE2 =<br>436 ms, TR: 5730 ms | TE = 73 ms<br>TR = 6500 ms | TE = 73 ms<br>TR = 6500 ms | TE = 73 ms<br>TR = 6500 ms | TE = 73 ms<br>TR = 6500 ms |
| Voxel size [mm<sup>3</sup>] | 0.6 × 0.6 × 0.6<br>0.8 × 0.8 × 0.8 | 0.8 × 0.8 × 0.8 | 1.0 × 1.0 × 1.0 | 0.8 × 0.8 × 0.8 | 0.5 × 0.5 × 2.0 | 0.5 × 0.5 × 2.0 | 0.5 × 0.5 × 2.0 | 0.5 × 0.5 × 2.0 |
| Magnetic field<br>strength [T] | 7 | 3 | 3 | 7 | 3 | 3 | 3 | 3 |
| Scanner<br>model/vendor | Philips Achieva<br>(Philips Healthcare,<br>Best, the<br>Netherlands) | Siemens Tim Trio<br>(Siemens<br>Healthineers,<br>Erlangen, Germany) | Siemens, GE, and<br>Philips (multi-vendor) | Siemens Magnetom<br>(Siemens<br>Healthineers,<br>Erlangen, Germany) | Siemens Skyra<br>(Siemens<br>Healthineers,<br>Erlangen, Germany) | Siemens Skyra<br>(Siemens<br>Healthineers,<br>Erlangen, Germany) | Siemens Skyra<br>(Siemens<br>Healthineers,<br>Erlangen, Germany) | Siemens Skyra<br>(Siemens<br>Healthineers,<br>Erlangen, Germany) |
| PVS burden |  |  |  |  |  |  |  |  |
| BG PVS volume<br>[ml] | 1.052<br>[0.889–1.195] | 0.263<br>[0.212–0.342] | 0.051<br>[0.022–0.099] | 0.920<br>[0.781–2.496] | 0.779<br>[0.592–1.255] | 0.587<br>[0.435–0.928] | 0.699<br>[0.459–0.992] | 2.388<br>[1.943–3.411] |
| Fractional BG PVS<br>volumes [%] | 1.767<br>[1.403–2.107] | 0.587<br>[0.416–0.611] | 0.117<br>[0.052–0.232] | 1.799<br>[1.459–4.536] | 1.865<br>[1.394–2.768] | 1.172<br>[0.945–1.839] | 1.585<br>[1.118–2.046] | 4.183<br>[3.185–5.685] |
| CSO PVS volume<br>[ml] | 7.163<br>[6.762–7.518] | 0.976<br>[0.545–1.686] | 0.900<br>[0.479–1.4305] | 6.713<br>[5.506–10.689] | 8.471<br>[5.854–9.552] | 6.853<br>[5.266–8.691] | 6.971<br>[4.275–7.397] | 21.672<br>[17.098–26.237] |
| Fractional CSO<br>PVS volumes [%] | 1.538<br>[1.517–1.7854] | 0.344<br>[0.192–0.652] | 0.387<br>[0.216–0.598] | 1.780<br>[1.378–2.699] | 3.027<br>[1.852–3.801] | 2.200<br>[1.678–2.822] | 2.446<br>[1.468–2.579] | 5.560<br>[4.175–7.258] |
| WMH burden |  |  |  |  |  |  |  |  |
| BG WMH volume<br>[ml] | No FLAIR imaging | No FLAIR imaging | 0.408<br>[0.103–0.878] | 0.075<br>[0.017–0.563] | No FLAIR imaging | 0.218<br>[0.017–0.837] | 0.293<br>[0.032–1.537] | 0.711<br>[0.405–1.153] |
| CSO WMH volume<br>[ml] | No FLAIR imaging | No FLAIR imaging | 3.364<br>[1.374–11.382] | 5.599<br>[3.827–6.735] | No FLAIR imaging | 1.510<br>[0.243–7.573] | 2.081<br>[0.155–9.819] | 5.871<br>[2.727–8.855] |

Reference annotations were generated by medical residents, neuroscientists, and experienced image analysts under the supervision of experienced neuroradiologists. Annotators manually delineated PVS using Mango or ITK-SNAP, following the STRIVE criteria (Duering et al., 2023). PVS were annotated on T1w scans for PCB, ADNI-3, and REMIND, and on T2w scans for EBBIVD, the HIM Study, MD-DARS, BB-DARS, and GlyM. Annotations covered every axial slice across the whole brain and were performed with the smallest available painting tool. Where available, FLAIR images were reviewed to distinguish PVS from WMH and exclude the latter from the reference annotations.

Under the DoRA-PVS Challenge protocol, inter-rater reliability in the basal ganglia (BG) ROI was a median Dice similarity coefficient (DSC) of 0.646 [IQR 0.598–0.651] and centreline DSC (clDice) of 0.684 [IQR 0.634–0.694]. Corresponding values in the centrum semiovale (CSO) ROI were 0.642 [IQR 0.624–0.659] for DSC and 0.687 [IQR 0.659–0.732] for clDice. These DSC values fall within the previously reported range of inter-rater agreement for PVS segmentation (Spijkerman et al., 2022). Intra-rater reliability was slightly higher: median BG DSC and clDice were 0.686 [IQR 0.664–0.724] and 0.749 [IQR 0.713–0.797], respectively, while the corresponding CSO ROI values were 0.668 [IQR 0.622–0.725] and 0.734 [IQR 0.674–0.758].

Ethical approval for each cohort was obtained from the relevant institutional review board. Further details are provided in the *Additional Information—Ethics approval section*. All participants provided written informed consent in accordance with the Declaration of Helsinki.

### 3.2 Regions of interest

The basal ganglia (BG) and centrum semiovale (CSO) ROIs were predefined by the DoRA-PVS Challenge 2026 and applied consistently to evaluate all submissions, including DRIPS. The ROI generation procedure was the following. Subject-specific basal ganglia (BG) and centrum semiovale (CSO) masks were derived from whole-brain anatomical labels generated from each T1w or T2w image using SynthSeg with the parcellation option enabled (Billot et al., 2023a). The BG ROI comprised the caudate nucleus, lentiform nucleus, thalamus, and internal and external capsules. As SynthSeg does not explicitly label the capsules, these were approximated independently in each hemisphere through morphological closing: the external capsule was defined as the white-matter compartment between the insular cortex and putamen, and the internal capsule as the space bounded by the caudate nucleus, putamen/pallidum, and thalamus. The CSO ROI comprised supratentorial white matter after exclusion of the BG ROI.

To address common segmentation errors around the ventricular atrium, choroid plexus, and posterior horns of the lateral ventricles, a dedicated exclusion template and masks (https://doi.org/10.7488/ds/7744) were registered to each participant’s native-space T1w image using combined linear and nonlinear SynthMorph transformations (Hoffmann et al., 2024). The resulting transforms were applied to the exclusion masks with nearest-neighbour interpolation. All refined ROI masks underwent visual inspection.

### 3.3 Evaluation metrics

Performance was assessed using threshold-free and threshold-dependent metrics. The threshold-free metric was the area under the precision-recall curve (AUPRC), which is well suited to highly imbalanced data (Maier-Hein et al., 2024) and allows for comparison with non-probabilistic, semi-automated methods whose performance depends strongly on the selected threshold. The threshold-dependent metric was the clDSC, which quantifies centreline overlap and is therefore well suited to thin, tubular structures (Maier-Hein et al., 2024). Since clDice depends on the binarisation threshold—and a fixed threshold of 0.5 is unlikely to be optimal across cohorts or appropriate for non-probabilistic methods—we also plot clDice across a range of thresholds.

### 3.4 Competing methods

We compared DRIPS with five previously published comparator methods evaluated within the DoRA-PVS Challenge 2026 framework (Dash et al., 2027): two non-probabilistic, non-machine learning methods—the Frangi filter (Frangi et al., 1998), and RORPO (Merveille et al., 2018, 2014)—and three deep learning methods. The deep learning methods were WAVM, whose two submissions (WAVM and WAVM2) extended the method of Pham et al. (Pham et al., 2026); GIN/IMN, which participated with SHIVAi (v3/T1-PVS) (Boutinaud et al., 2021); and VICOROBIGR, which submitted an nnU-Net-based approach (Isensee et al., 2021) trained on data from the VALDO Challenge (Sudre et al., 2024) and a private dataset.

## 4 Results

### 4.1 Ablation study

As part of the ablation study, we examined the effects of the intensity-mixing coefficient (*α_c_*) during PVS generation, simulated motion artefacts, the Laplacian prior, and voxel resampling (*p*_iso_), and network dimensionality (2D vs 3D) on DRIPS performance.

#### 4.1.1 Effect of the intensity-mixing coefficient ***α_c_***

The parameter *α_c_* is a component-specific intensity-mixing coefficient that controls the visibility, or contrast, of each simulated PVS relative to its surrounding tissue. Lower values retain more of the background-tissue intensity, producing less salient PVS. To assess the effect of contrast variability, we compared a model trained with a fixed mixing coefficient (*α*_c_ = 1) with one trained using a coefficient sampled over a wider range (*α*_c_∼*<u>U</u>*(0.51, 2.00)) (Figure 4). In the BG ROI, the fixed-coefficient model achieved higher AUPRC (0.524 [IQR 0.454–0.615] vs 0.491 [IQR 0.407–0.584], P = 0.016), but clDice did not differ significantly between models (0.484 [IQR 0.425–0.545] vs 0.473 [IQR 0.429–0.502], P = 0.844). On the other hand, in the CSO ROI, sampling *α_c_* over the wider range improved both AUPRC (0.423 [IQR 0.294–0.585] vs 0.365 [IQR 0.276–0.457], P = 0.008) and clDice (0.425 [IQR 0.322–0.541] vs 0.355 [IQR 0.297–0.467], P = 0.008).

**Figure 4.**
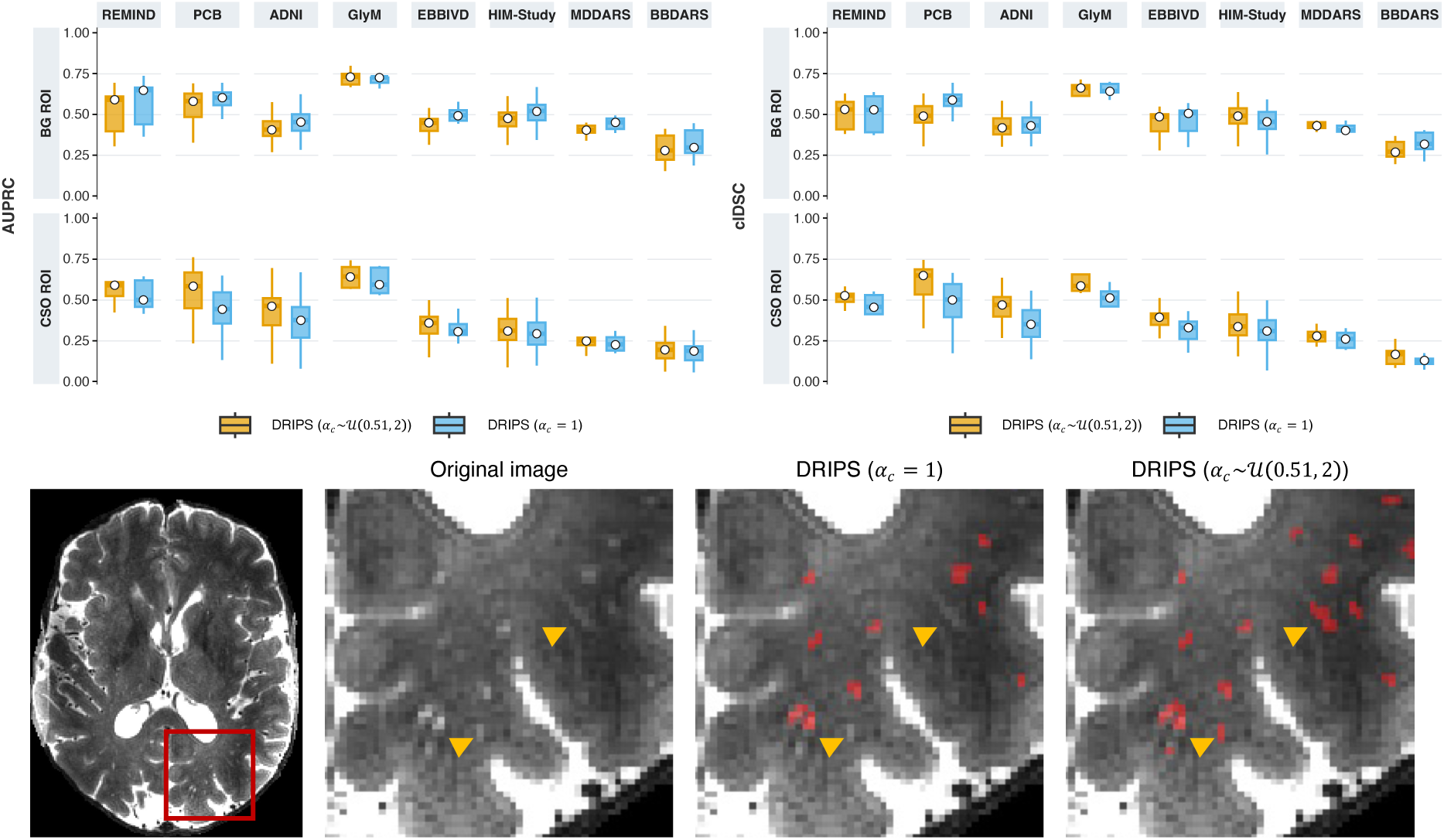
Varying the mixing between the synthetic PVS intensity and that of the surrounding tissue during image synthesis generally improved the identification of subtle PVS. The largest gains were observed in the CSO ROI, where contrast variability yielded significantly higher AUPRC and clDice than a fixed mixing coefficient. The images at the bottom illustrate this effect: subtle PVS (yellow arrow) were missed by the model trained with a fixed coefficient but identified when the model was trained with a range of mixing coefficients. In the BG ROI, a fixed coefficient yielded higher AUPRC; however, it had little effect on the final spatial overlap of PVS segmentations.

#### 4.1.2 Effect of motion artefact simulation

To assess the effect of simulated motion artefacts on DRIPS performance, we compared models trained with and without motion simulation (Figure 5). Motion simulation had no significant effect on clDice in either ROI: 0.473 [IQR 0.429–0.502] vs 0.444 [IQR 0.390–0.502] in the BG ROI (P = 0.383), and 0.425 [IQR 0.322–0.541] vs 0.369 [IQR 0.320–0.436] in the CSO ROI (P = 0.078). However, it significantly improved CSO AUPRC (0.423 [IQR 0.294–0.585] vs 0.367 [IQR 0.287–0.440], P = 0.016). In the BG ROI, AUPRC did not differ significantly between models (0.491 [IQR 0.407–0.584] vs 0.503 [IQR 0.458–0.584], P = 0.250).

**Figure 5.**
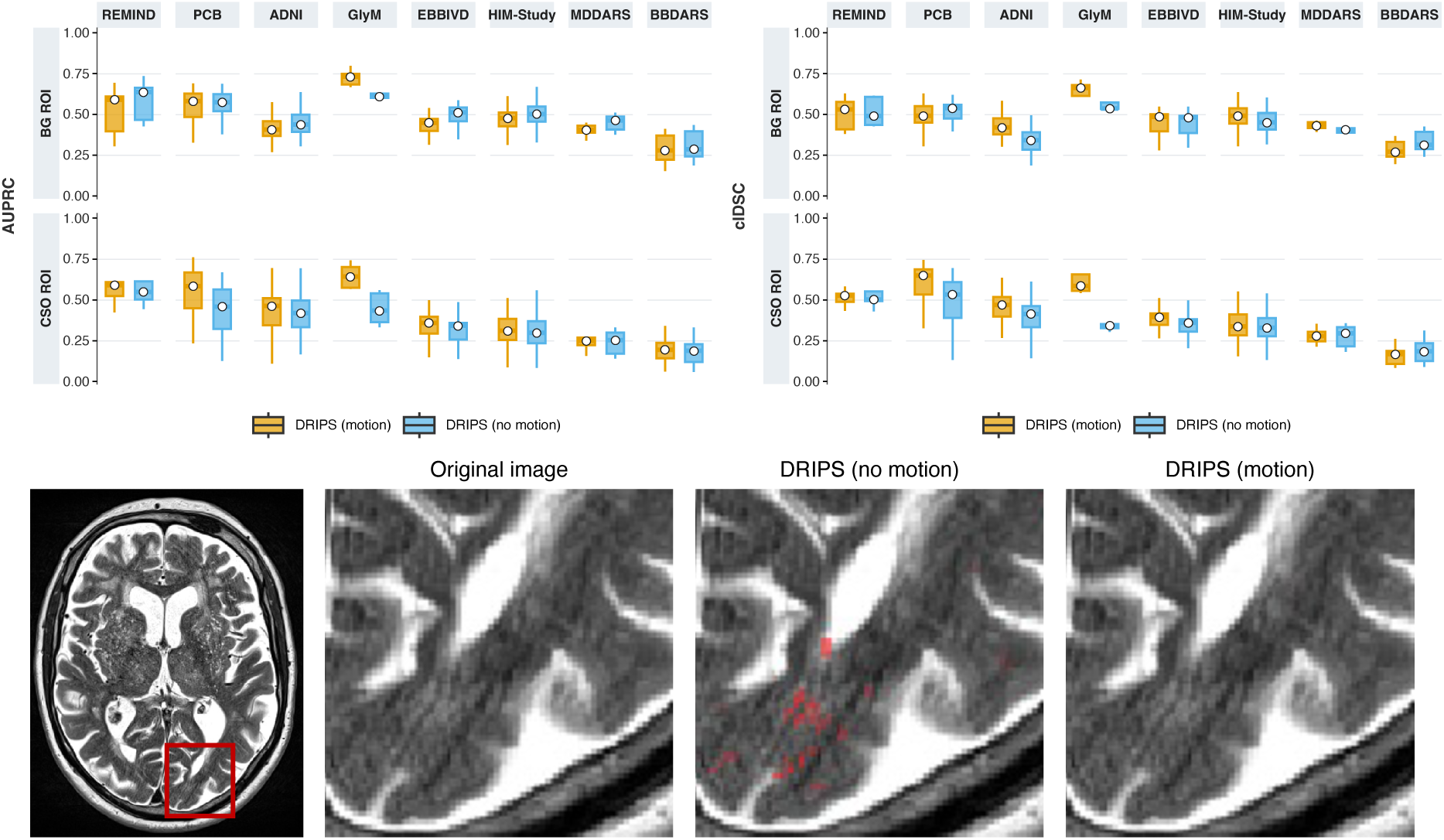
Simulating motion artefacts during image synthesis produced a more conservative model that was less prone to labelling motion-related signal as PVS. Motion simulation improved CSO AUPRC and clDice, i.e., it helped discriminate better between true PVS and motion-related signal. The example at the bottom illustrates this effect: the model trained without motion simulation incorrectly segmented visible motion artefacts as PVS, whereas the motion-trained model was more robust to these artefacts. For this case, clDice increased from 0.376 without motion simulation to 0.396 with motion simulation. The BG AUPRC was however lower, suggesting that the more conservative behaviour may have reduced the model’s confidence in distinguishing some true PVS from background.

#### 4.1.3 Effect of the Laplacian prior

To assess the contribution of the Laplacian prior, which emphasises local intensity changes compatible with PVS, we compared DRIPS with and without this component (Figure 6). Adding the Laplacian prior significantly improved the ability of the model to distinguish PVS from other structures, increasing BG AUPRC from 0.431 [IQR 0.346–0.567] to 0.491 [IQR 0.407–0.584] (P = 0.008) and CSO AUPRC from 0.356 [IQR 0.250–0.524] to 0.423 [IQR 0.294–0.585] (P = 0.008). It had, however, no significant effect on clDice in either region: 0.473 [IQR 0.429–0.502] versus 0.478 [IQR 0.423–0.561] in the BG ROI (P = 1.000), and 0.425 [IQR 0.322–0.541] versus 0.396 [IQR 0.311–0.548] in the CSO ROI (P = 0.195).

**Figure 6.**
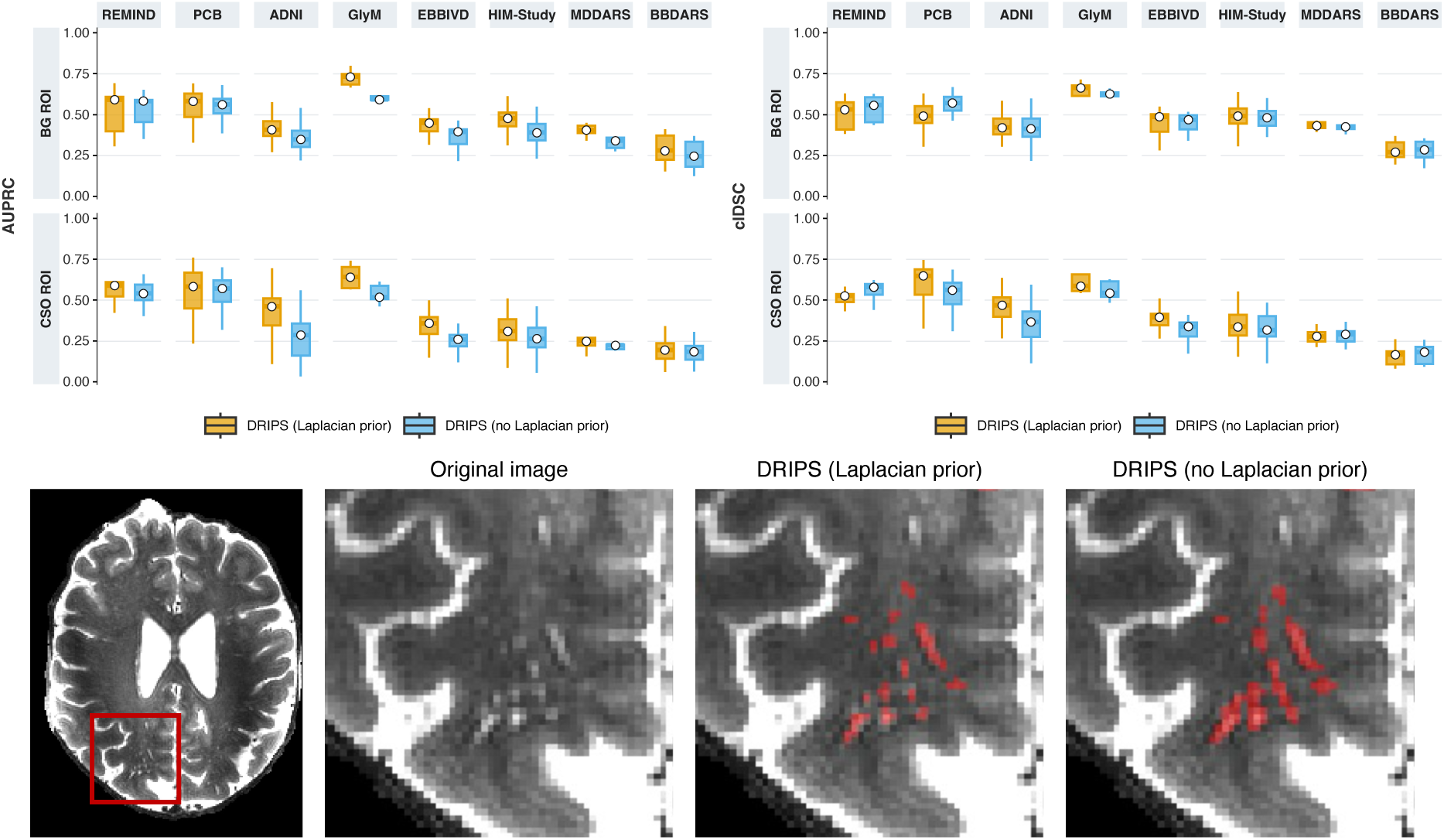
The Laplacian prior improved the model’s ability to distinguish PVS from background across thresholds, without a measurable change in clDice. This pattern is evident in the upper plots: AUPRC was higher in many cohorts when the Laplacian prior was used, whereas clDice remained similar. The lower example illustrates why. The main effect of the Laplacian prior was to refine PVS delineation: without it, neighbouring individual PVS were often merged into a single contiguous segmentation, whereas the Laplacian-prior model more clearly separated them as distinct structures.

**Figure 7.**
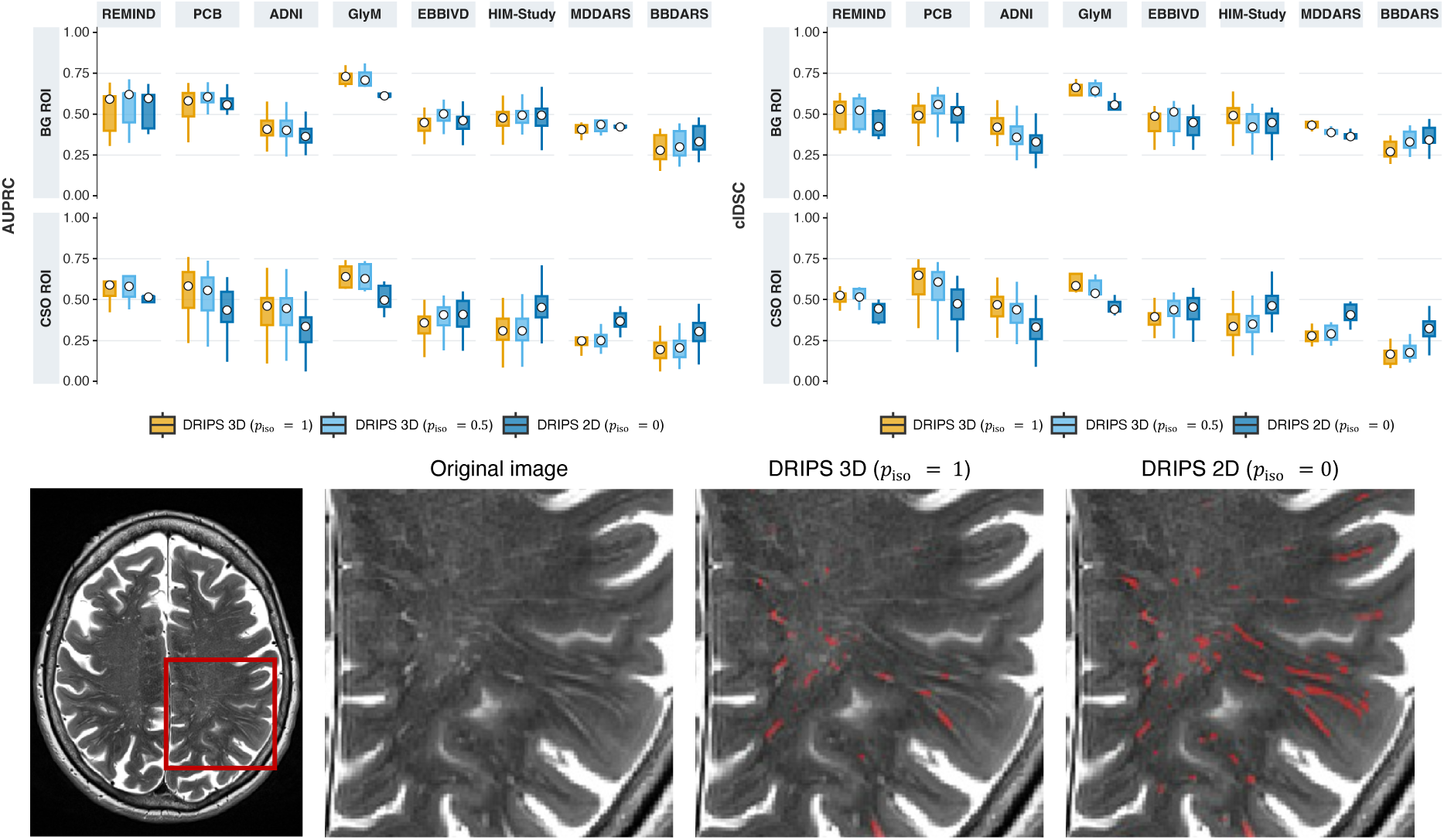
Using a 2D network for anisotropic scans and a 3D network for isotropic scans is the better choice. The upper plots show that the 3D network generally performed better in isotropic cohorts (REMIND, PCB, ADNI, and GlyM), but performed poorly in anisotropic cohorts (EBBIVD, HIM-Study, MD-DARS, and BB-DARS). Training a single 3D model on both isotropic and anisotropic images (e.g., when *p*_iso_ = 0.5) did not adequately address this limitation. These findings support selecting the network dimensionality according to image resolution: 3D for isotropic and 2D for anisotropic acquisitions. The lower example illustrates this issue. Although the 3D model detected parts of the visible PVS on this case from BBDARS, it failed to segment their full extent, whereas the 2D model produced a more complete delineation (CSO clDice: 0.261 vs 0.462).

#### 4.1.4 Effect of voxel resampling ***p***_iso_ and network dimensionality

To assess whether network dimensionality should be adapted to image resolution, we compared a 2D model trained on anisotropic images (*p*_iso_ = 0) with 3D models trained on exclusively isotropic images (*p*_iso_ = 1) or on a mixture of isotropic and anisotropic images (*p*_iso_ = 0.5) (Figure 7). Across the four isotropic cohorts, the 3D model trained exclusively on isotropic images (*p*_iso_ = 1) achieved the strongest CSO performance, with AUPRC of 0.569 [IQR 0.553–0.603] and clDice of 0.558 [IQR 0.512–0.602], compared with 0.554 [IQR 0.530–0.593] and 0.525 [IQR 0.497–0.556] for the mixed 3D model (*p*_iso_ = 0.5), and 0.447 [IQR 0.412–0.502] and 0.423 [IQR 0.413–0.453] for the 2D model. In contrast, across the four anisotropic cohorts, the 2D model achieved substantially higher CSO AUPRC (0.384 [IQR 0.353–0.419]) and clDice (0.410 [IQR 0.387–0.450]) than either the mixed 3D model (0.292 [IQR 0.239–0.333] and 0.313 [IQR 0.262–0.371]) or the isotropic-only 3D model (0.276 [IQR 0.235–0.320] and 0.293 [IQR 0.250–0.350]). Differences in the BG ROI were more modest. Thus, mixing isotropic and anisotropic images during 3D training improved performance relative to isotropic-only 3D training on anisotropic data, but did not match a 2D model specialised for anisotropic acquisitions.

#### 4.1.5 Final DRIPS configuration

Taken together, the ablation analyses support the following DRIPS configuration: a 3D network trained exclusively on isotropic scans for isotropic acquisitions, and a 2D network for anisotropic acquisitions, both incorporating a Laplacian prior, a variable rather than fixed intensity-mixing coefficient *α_c_*, and simulated motion artefacts during synthesis. The results reported in Sections 4.2 and 4.3 correspond to those obtained using this final configuration.

### 4.2 Comparison against competing methods

DRIPS achieved the highest overall performance in both ROIs across the eight cohorts (Figure 8 and Figure 9). DRIPS outperformed the second-best method in BG AUPRC (0.503 [IQR 0.419–0.584] vs 0.356 [IQR 0.327–0.402] for SHIVAi; mean difference: 0.146, P = 0.008), CSO AUPRC (0.476 [IQR 0.398–0.585] vs 0.351 [IQR 0.253–0.425] for WAVM2; mean difference: 0.125, P = 0.008), BG clDice (0.463 [IQR 0.406–0.502] vs 0.351 [IQR 0.277–0.400] for SHIVAi; mean difference: 0.112, P = 0.016), and CSO clDice (0.484 [IQR 0.436–0.541] vs 0.298 [IQR 0.205–0.333] for WAVM2; mean difference: 0.186, P = 0.008). DRIPS was significantly outperformed only for BG AUPRC in ADNI, where Frangi achieved a higher value (0.429 vs 0.408, P = 0.015). For BG clDice, DRIPS showed comparable performance to SHIVAi in ADNI (0.420 vs 0.426, P = 0.877) and Frangi in MD-DARS (0.364 vs 0.385, P = 0.313).

**Figure 8.**
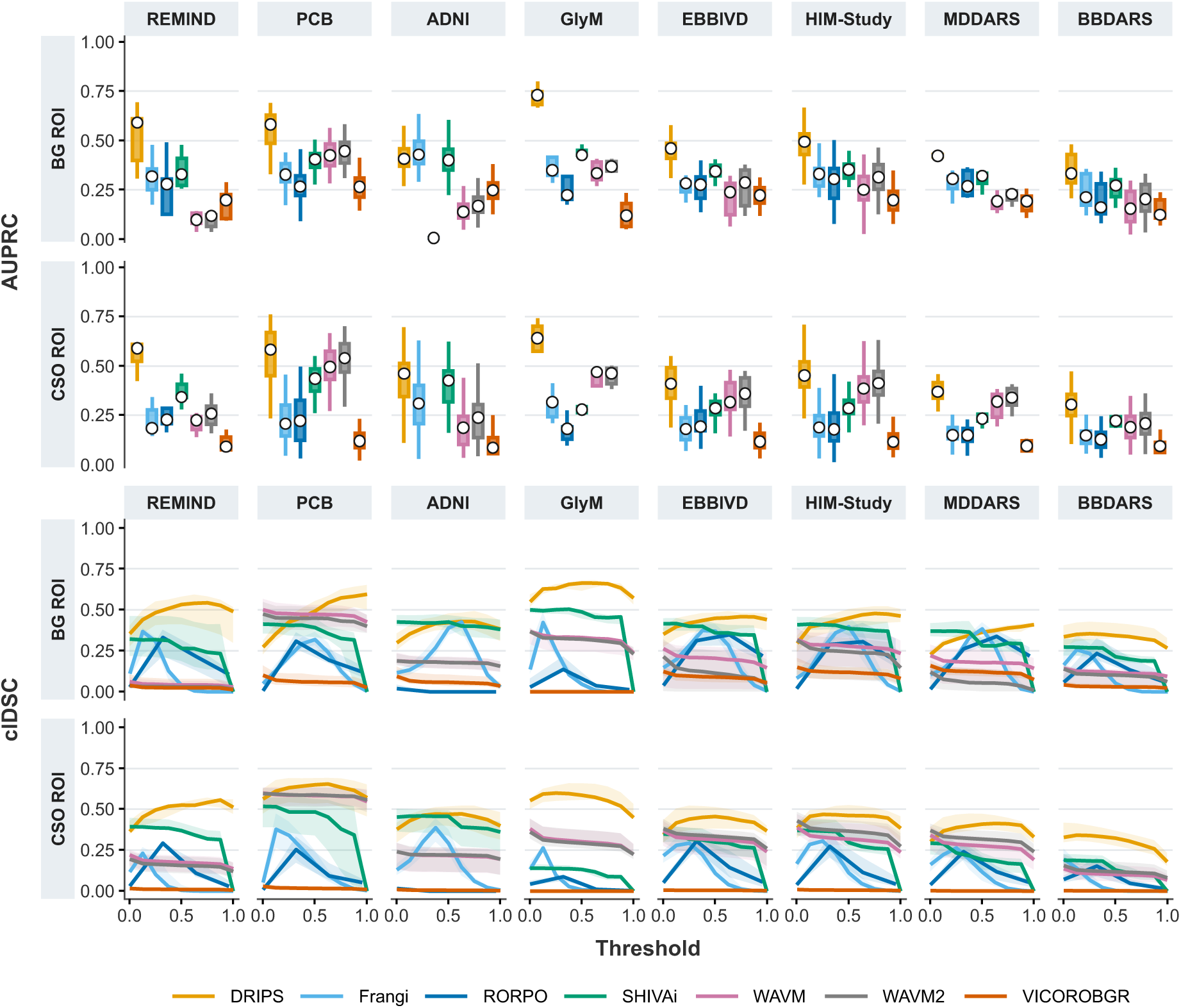
Performance of DRIPS and competing methods across test cohorts. PVS segmentation was evaluated in the basal ganglia (BG ROI) and centrum semiovale (CSO ROI) using the area under the precision–recall curve (AUPRC) and centreline Dice similarity coefficient (clDice). AUPRC summarises discrimination across the full range of response values, whereas clDice quantifies the overlap and continuity of the segmented PVS centrelines. Because PVS visibility—and thus the sensitivity of the manual reference delineations—varies with image-acquisition protocol, clDice is shown across thresholds from 0 to 1 rather than at a single operating point. To place the non-probabilistic Frangi and RORPO outputs on the same threshold axis and enable their visualisation, their responses were transformed. For the Frangi filter, the original vesselness response was transformed as *frangi* = log(10^6^* · *frangi + 1) / 6.* For RORPO, the normalised threshold was *rorpo = rorpo / 50*. Consequently, a normalised threshold of 0.5 corresponds to an original Frangi response of 1.91×10^-5^ and a RORPO threshold of 25.

**Figure 9.**
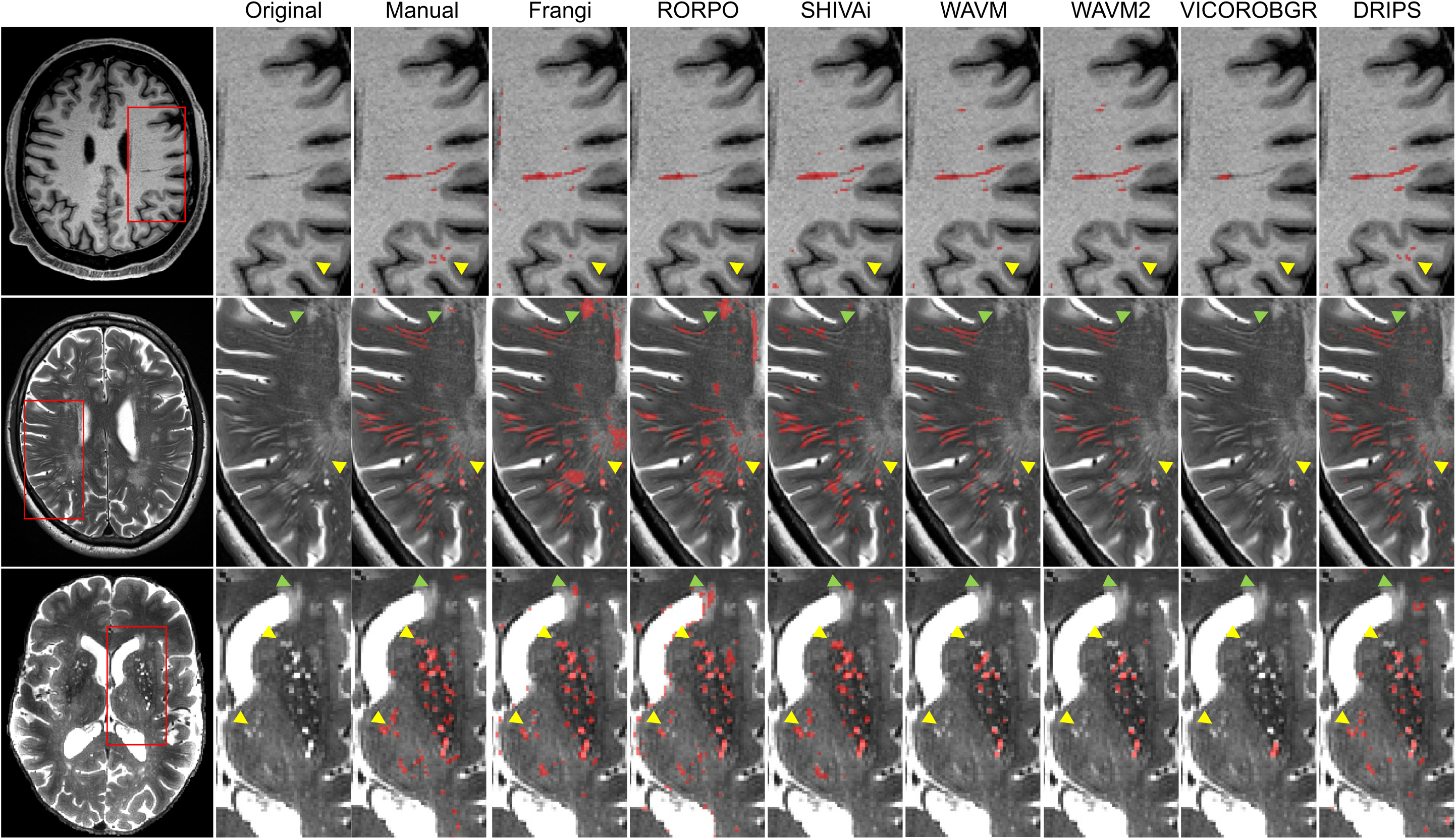
Representative segmentation maps from DRIPS and competing methods. These cases are an isotropic PCB scan (top), an anisotropic HIM-Study scan (middle), and an isotropic GlyM scan (bottom). The “manual” column shows the reference segmentation. Yellow arrows highlight PVS missed by multiple methods, while green arrows indicate white matter hyperintensities incorrectly segmented as PVS.

Threshold-clDice curves indicated distinct operating characteristics across methods (Figure 8). WAVM, WAVM2, and VICOROBGR achieved their highest clDice at the lowest sampled threshold, while SHIVAi peaked at the lowest thresholds. Frangi and RORPO, in contrast, achieved their highest performance over narrow threshold ranges, which varied from one cohort to another. DRIPS showed relatively stable performance across intermediate thresholds. In the BG ROI, median clDice was 0.449 at the threshold of 0.5 and reached a maximum of 0.468 at 0.625, remaining within 10% of this maximum between thresholds of 0.375 and 1. In the CSO ROI, clDice at 0.5 was 0.481, closely matching the maximum of 0.482 at 0.375, and remained within 10% of its maximum between 0.125 and 0.875. The BG ROI in PCB was an exception: clDice continued to increase up to the highest sampled threshold, suggesting that DRIPS may have identified more voxels as PVS than were represented in the manual delineations.

### 4.3 WMH and PVS segmentation

We examined whether WMH burden was associated with segmentation performance using a linear mixed-effects model of AUPRC with method-specific WMH-volume effects, adjusted for reference PVS volume, ROI, and cohort, and with a random intercept for case (Figure 10). DRIPS AUPRC was not associated with WMH volume (P=0.254). The same was observed for RORPO, SHIVAi, WAVM, WAVM2, and VICOROBGR (all P ≥ 0.070). In contrast, Frangi performance decreased as WMH burden increased: for otherwise comparable regions, a tenfold increase in WMH volume (e.g., from 0.1 to 1 mL) was associated with an estimated 0.032-point lower AUPRC (B =-0.032 [95% CI-0.046,-0.018], P<0.001).

**Figure 10.**
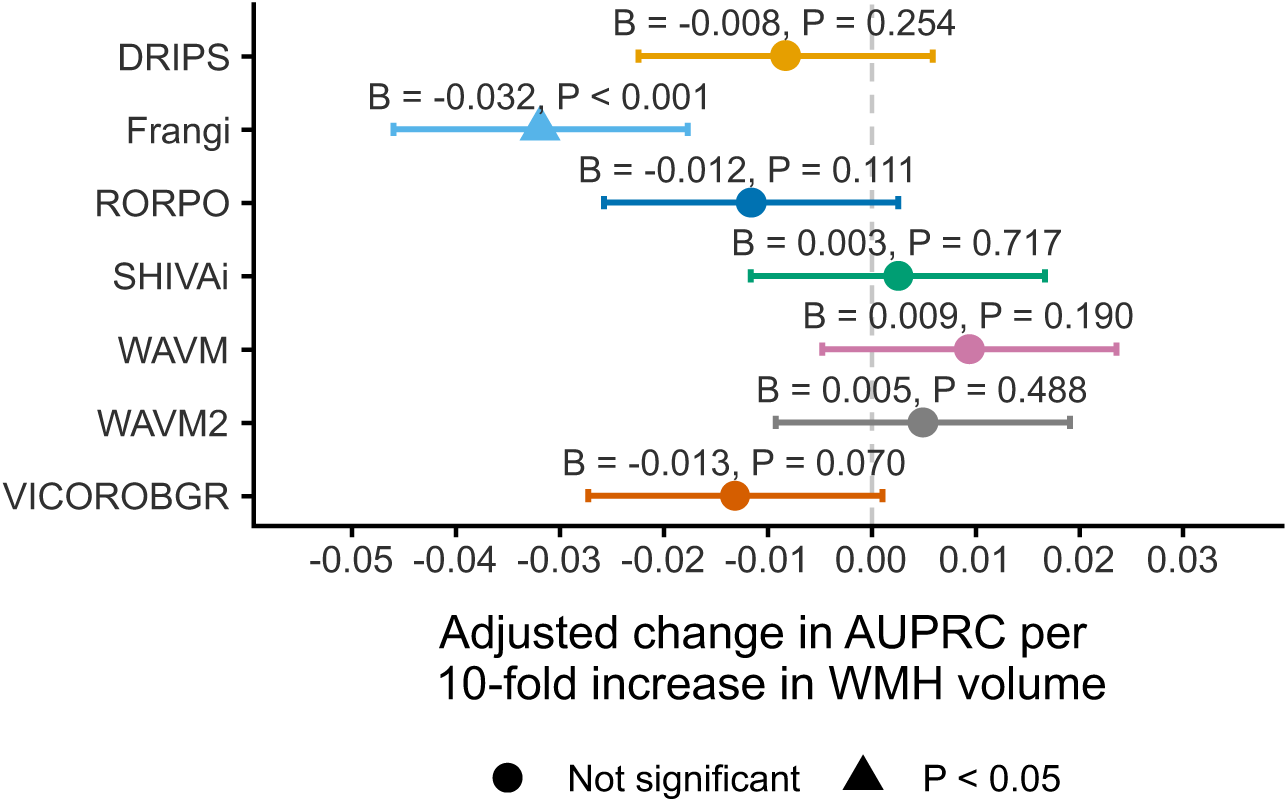
Association between segmentation performance and WMH burden after accounting for PVS volume. Method-specific estimates show the adjusted change in AUPRC per 10-fold increase in region-matched WMH volume from a linear mixed-effects model controlling for reference PVS volume, ROI, and cohort, with a random intercept for case. Only the Frangi filter showed lower performance with increasing WMH volume.

## 5 Discussion

In recent years, deep learning has become the dominant paradigm for PVS segmentation (Waymont et al., 2024) and for medical image analysis more broadly, because it can learn task-relevant representations directly from data. and medical image analysis more broadly, owing to its ability to learn task-relevant representations directly from data. This is partly supported by our evaluation: despite being assessed out of sample, deep-learning methods generally outperformed non-probabilistic approaches across the evaluated datasets. However, deep learning models are data-hungry. Large, heterogeneous, manually annotated datasets remain scarce in medical imaging, and training on limited or homogeneous data can encourage shortcut learning rather than robust anatomical representations. This was also evident in our evaluation: performance was lower on isotropic 7 T than on 3 T scans, with the principal failure mode being an inability to reliably identify PVS (e.g., CSO PVS in GlyM for SHIVAi and PVS in REMIND for WAVM). Although models can evidently be retrained when new failure modes emerge, relying on manual annotation and dataset-specific fine-tuning every time this happens is neither scalable nor sustainable long-term.

Against this backdrop, we present DRIPS, a PVS segmentation framework trained exclusively on synthetic data that nevertheless accurately segments PVS in real-world imaging data. Across eight independent cohorts, we show that DRIPS (i) segments PVS in isotropic and anisotropic T1- and T2-weighted images accurately; (ii) generally outperforms non-machine-learning and machine-learning-based approaches; and (iii) provides PVS estimates independently of WMH burden. These findings position DRIPS as a robust and versatile framework for PVS segmentation.

### 5.1 DRIPS works without real PVS annotations

A key benefit of domain randomisation is its ability to generate diverse image-label pairs that, when appropriately designed, can support generalisation to real world data. This strategy has been effective when target structures are already available as label maps, including anatomical regions, tissue classes, and existing lesion masks (Billot et al., 2023a; Chalcroft et al., 2025; Hoopes et al., 2022; Laso et al., 2024; Valabregue et al., 2026). A more challenging scenario arises when voxel-level lesion annotations are unavailable altogether. Here, we show that real PVS annotations are not required for training, provided that key radiological characteristics are incorporated during synthesis, including their tubular morphology, distribution in white matter and subcortical structures, and orientation towards the lateral ventricles. As other cerebral small vessel disease markers similarly have characteristic morphology, anatomical distribution, and intensity polarity, strategies similar to the one presented here may also be applicable to their segmentation, for example, for cortical superficial siderosis or cerebral microbleeds (Cao et al., 2026).

### 5.2 DRIPS segments PVS accurately on real MRI data

We compared DRIPS with five previously published methods submitted to the DoRA-PVS Challenge 2026 using 185 fully manually annotated cases from eight cohorts, encompassing diverse clinical conditions and a broad range of scanners and acquisition protocols. Despite this substantial heterogeneity, DRIPS outperformed the comparator methods in most cases for both AUPRC and clDice, supporting its robustness relative to current state-of-the-art approaches.

PVS segmentation nevertheless remains challenging. At a threshold of 0.5, mean cross-cohort clDice was 0.463 [IQR 0.406–0.502] in the BG ROI and 0.484 [IQR 0.436–0.541] in the CSO ROI. By comparison, inter-rater clDice in the same regions reached 0.684 [IQR 0.634–0.694] and 0.687 [IQR 0.659–0.732], respectively. Thus, even the best-performing automated method remained approximately 0.2 clDice points below human inter-rater agreement, indicating there is a gap for further methodological improvement, esp. for highly anisotropic acquisitions.

### 5.3 Threshold selection and the need for quality control

Threshold sweeps confirmed the common consensus that the optimal operating point generally varies across cohorts for all methods. However, the threshold that maximised performance was strongly method-specific. Frangi and RORPO performed well only over narrow threshold ranges, making their transfer across scanners and protocols challenging since modest changes in the selected cut-off can substantially alter the resulting segmentation.

For SHIVAi, WAVM, and VICOROBGR, the highest clDice was achieved at the lowest sampled thresholds, consistent with many genuine PVS receiving low response values and therefore being excluded at more stringent cut-offs. This pattern may partly reflect their use of resampling to a fixed reference voxel spacing before inference, followed by resampling of predictions back to native space. In high-resolution acquisitions, the initial downsampling can blur or remove subtle PVS, and upsampling cannot restore this lost information. Their achievable sensitivity may therefore be constrained for images with voxels sizes far from the fixed reference voxel spacing.

The DRIPS curve resembled a plateau rather than a peak. In both ROIs, the prespecified threshold of 0.5 yielded clDice close to the optimum across the sweep, supporting the pragmatic use of a common threshold across heterogeneous cohorts and reducing the need for dataset-specific tuning. PCB was the exception: in the BG ROI, agreement between DRIPS and the manual annotations was highest at the highest sampled thresholds, i.e. DRIPS was over-sensitive compared to the human raters in this cohort.

Regardless of whether a method is machine-learning-based, the practical implication is straightforward: segmentations should be visually quality-controlled before large-scale deployment to verify that the selected threshold is appropriate for the target data.

### 5.4 DRIPS is robust to the presence of WMH

WMHs have long been considered a confounder for PVS segmentation: on a single structural image, they can either have signal profiles similar to those of PVS or conceal small PVS altogether (Waymont et al., 2024). Our results suggest that this vulnerability is however not inevitable. After accounting for PVS volume, increasing WMH burden was associated with poorer performance only for the Frangi filter, not for the other evaluated methods. This points to a specific limitation of vesselness filtering and further supports the common practice of combining Frangi responses with complementary contrasts, such as FLAIR, to distinguish WMHs from PVS. Although the sample was limited, our findings indicate that PVS segmentation can be made substantially less sensitive to co-occurring lesions—a critical prerequisite for unbiased analyses in cerebrovascular disease.

### 5.5 Limitations and future work

Despite the demonstrated advantages of our approach, five limitations merit consideration. First, we modelled PVS as tortuous tubular structures distributed throughout the brain. While effective for training and segmentation, this abstraction oversimplifies their biology. *In vivo*, PVS are closely aligned with the cerebral vasculature, following the trajectories of arterioles, capillaries, and venules, with their orientation, calibre, and spatial density shaped by vascular anatomy, regional blood supply, and vessel tortuosity. Second, we assumed a predominant orientation towards the lateral ventricles. This is a reasonable approximation for PVS in the centrum semiovale, which often follow medullary arteries radiating to the ventricles, but it does not hold in other regions—for instance, PVS surrounding the lenticulostriate arteries in the basal ganglia, which typically run perpendicular to the axial plane. Looking ahead, these limitations highlight an opportunity: conditioning PVS generation on vascular maps could produce more physiologically plausible simulations, improving anatomical fidelity and reducing false positives in regions where tubular structures occur independently of vessels. Third, DRIPS used a conventional 3D U-Net for isotropic acquisitions and a 2D U-Net for anisotropic acquisitions. We did not evaluate more advanced network architectures. Although this is a limitation, architectural optimisation was not the primary aim of this study. Instead, we sought to establish whether the DRIPS framework could achieve accurate and robust PVS segmentation across cohorts, health conditions, and imaging settings. Future work should assess whether more sophisticated architectures, including transformers and models with anisotropic convolutions, can further improve performance. Fourth, although our evaluation included data from multiple individuals across five cohorts spanning a wide range of conditions, our assessment remains limited by the imaging protocols included in this study. As part of our future work, we plan to include patients spanning a broader range of disease severities—from very mild to advanced—scanned using multiple imaging sequences, and to conduct longitudinal assessments to evaluate the method’s ability to track changes in PVS over extended periods of time. Fifth, we simulated only a subset of possible artefacts and did not systematically assess whether alternative simulation strategies could further improve performance. Future work should evaluate additional artefact-generation approaches, e.g., 10.7488/ds/8099.

## 6 Conclusion

We present DRIPS, the first domain-randomisation framework for accurate out-of-sample PVS segmentation. Without manual PVS annotations, retraining, or fine-tuning, DRIPS accurately segmented PVS in 3 T and 7 T T1- and T2-weighted images across isotropic and anisotropic resolutions, and outperformed five competing methods across most scenarios. Unlike competing methods, its performance was not associated with total WMH burden. These findings demonstrate that DRIPS enables robust PVS quantification across diverse imaging settings and patient populations, supporting more accessible and reliable use in research and clinical applications.

## Acknowledgements

This work was supported by the German Centre for Neurodegenerative Diseases (DZNE; BN012), the German Research Foundation (DFG; 425899996/CRC 1436, projects A05, B02, B04 and C01; 362321501/RTG 2413 SynAGE; MA 9235/3-1/SCHR 1418/5-1 (501214112)), the Deutsche Alzheimer Gesellschaft e.V. and Förderstiftung Dierichs (MD-DARS and BB-DARS), the Galen and Hilary Weston Foundation/Weston Brain Institute (UB190097), and the Row Fogo Charitable Trust/Row Fogo Centre for Research into Ageing and the Brain (AD.ROW4.35, BRO-D.FID3668413). PCB was partly funded by IZKF Jena and BMBF start-up funding for the German Centre for Mental Health (DZPG; BMBF 01EE2305D, JE2/TP5). The HIM-Study was supported by the European Regional Development Fund (EFRE; ZS/2024/02/184014 and ZS/2024/05/187256) and the Polycarp-Leporin-Program (PLP23/5). REMIND is funded by Stichting Woelse Waard and Amsterdam Neuroscience (proof of concept grant). Funders had no role in study design, data collection, analysis, interpretation, or manuscript preparation. Data collection and sharing for the Alzheimer’s Disease Neuroimaging Initiative (ADNI) is funded by the National Institute on Aging (National Institutes of Health Grant U19 AG024904). The grantee organization is the Northern California Institute for Research and Education. In the past, ADNI has also received funding from the National Institute of Biomedical Imaging and Bioengineering, the Canadian Institutes of Health Research, and private sector contributions through the Foundation for the National Institutes of Health (FNIH) including generous contributions from the following: AbbVie, Alzheimer’s Association; Alzheimer’s Drug Discovery Foundation; Araclon Biotech; BioClinica, Inc.; Biogen; Bristol-Myers Squibb Company; CereSpir, Inc.; Cogstate; Eisai Inc.; Elan Pharmaceuticals, Inc.; Eli Lilly and Company; EuroImmun; F. Hoffmann-La Roche Ltd and its affiliated company Genentech, Inc.; Fujirebio; GE Healthcare; IXICO Ltd.; Janssen Alzheimer Immunotherapy Research & Development, LLC.; Johnson & Johnson Pharmaceutical Research &Development LLC.; Lumosity; Lundbeck; Merck & Co., Inc.; Meso Scale Diagnostics, LLC.; NeuroRx Research; Neurotrack Technologies; Novartis Pharmaceuticals Corporation; Pfizer Inc.; Piramal Imaging; Servier; Takeda Pharmaceutical Company; and Transition Therapeutics. The funding bodies played no role in the design of the study or collection, analysis, or interpretation of data or in writing the manuscript.

## CRediT authorship contribution statement

Conceptualisation: L.B., M.Di., G.Z., J.B.; Methodology: L.B., M.Di., J.B.; Software: L.B., M.Di., J.B.; Validation: L.B., M.Di., J.B.; Formal analysis: L.B., M.Di., J.B.; Investigation: L.B., M.Di., J.B.; Resources: H.M, K.N, M.P, C.B., H.T.M., E.F., S.T., D.T., B.B., T.R., P.A.R, A.S., N.O., C.G., M.W., P.A., D.B., C.P., Y.L., P.M., R.B.D, S.S., E.D., G.Z., J.B. Data Curation: L.B., J.B.; Writing - Original Draft: L.B., J.B.; Writing - Review & Editing: All authors; Visualisation: L.B., J.B.; Supervision: J.B.; Project administration: J.B..

## Data availability statement

The MRI data analysed in this study cannot be made publicly available because they contain sensitive human participant data. Unrestricted deposition is precluded by confidentiality requirements and the informed-consent terms under which the data were collected. Researchers who wish to evaluate their algorithms on the datasets used in this work are invited to participate in the DoRA-PVS Challenge (https://www.synapse.org/Synapse:syn72039291/wiki/636166).

To support reproducibility and consistent deployment across systems, we provide a ready-to-use Docker image for DRIPS at https://github.com/MIA-FAU/DRIPS.

## Ethics approval and consent to participate

All participants gave written informed consent to participate in the study. Ethical approval was obtained from the Ethics Committees of University Hospital Magdeburg for the EBBIVD (93/17), HIM (82/22), MDDARS (93/17), and BBDARS (28/16) cohorts; Jena University Medical School for the PCB cohort (2021-2158_1-BO); Maastricht University Medical Center, the Netherlands, for the GlyM cohort (NL72269.068.19); and Amsterdam University Medical Centre for the REMIND cohort (2020.443). For ADNI-3, approval was obtained from the Institutional Review Boards or ethics committees at all participating centres before participant enrolment. The authors assert that all procedures contributing to this work comply with the ethical standards of the relevant national and institutional committees on human experimentation and with the Declaration of Helsinki and its later amendments.

## Declaration of generative AI and AI-assisted technologies in the manuscript preparation process

During the preparation of this work, the authors used ChatGPT (v5.6 Terra) for language and grammar editing. The authors subsequently reviewed and revised the content as needed and take full responsibility for the final published article.

