## Supplementary Material for "DRIPS: Domain Randomisation for Image-based Perivascular space Segmentation"

**Supplementary Table 1.** Parameter used during label and image synthesis and their corresponding values.

| Parameter | Value |
|---|---|
| $\theta_{\mathrm{min}}$ | -15 |
| $\theta_{\mathrm{max}}$ | 15 |
| $tr_{\mathrm{min}}$ | 0 |
| $tr_{\mathrm{max}}$ | 0 |
| $p_{\mathrm{iso}}$ | 1 for 3D; 0 for 2D |
| $r_{\mathrm{iso,min}}$ | 0.5 |
| $r_{\mathrm{iso,max}}$ | 1.0 |
| $r_{\perp,\mathrm{min}}$ | 1.1 |
| $r_{\perp,\mathrm{max}}$ | 4.0 |
| $r_{\|\|,\mathrm{min}}$ | 0.5 |
| $r_{\|\|,\mathrm{max}}$ | 1.0 |
| $C_{\mathrm{min}}$ | 100 |
| $C_{\mathrm{max}}$ | 500 |
| $L_{\mathrm{min}}$ | 3 |
| $L_{\mathrm{max}}$ | 30 |
| $\alpha_{\mathrm{c,min}}$ | 0.51 |
| $\alpha_{\mathrm{c,max}}$ | 2.00 |
| $b_B$ | 0.7 |
| $R_{\theta\mathrm{min}}$ | -15 |
| $R_{\theta\mathrm{max}}$ | 15 |
| $pr_{\mathrm{min}}$ | 0.5 |
| $pr_{\mathrm{max}}$ | 1.0 |
| $\mathrm{SNR}_{\mathrm{min}}$ | 20 |
| $\mathrm{SNR}_{\mathrm{max}}$ | 50 |
| $\gamma_{min}$ | 0.67 |
| $\gamma_{max}$ | 1.5 |
| $\alpha_{\mathrm{F}}$ | 0.75 |
| $\gamma_{\mathrm{F}}$ | 1.5 |
| $\alpha_T$ | 0.1 |
| $\beta_T$ | 0.9 |
